# The impact of botanical fermented foods on obesity, metabolic syndrome and type 2 diabetes: a systematic review of randomised controlled trials

**DOI:** 10.1101/2022.12.10.22283002

**Authors:** Miin Chan, Nadja Larsen, Helen Baxter, Lene Jespersen, Elif I Ekinci, Kate Howell

**Affiliations:** School of Agriculture and Food, University of Melbourne, Melbourne, Victoria, Australia; Department of Food Science, University of Copenhagen, Frederiksberg, Denmark; Austin Health Science Library, Austin Health, Heidelberg, Victoria, Australia; The Australian Centre for Accelerating Diabetes Innovations (ACADI), Melbourne Medical School, The University of Melbourne and Department of Endocrinology Austin Health

## Abstract

**Objective:** To assess whether botanical fermented food (BFF) consumption has an impact on cardiometabolic biomarkers or gut microbiota in adults with obesity, metabolic syndrome (MetS) or type 2 diabetes mellitus (T2DM).

**Design:** Systematic review

**Data sources:** Embase, MEDLINE, Cochrane CENTRAL and Google Scholar were searched with no language limits, from inception to August 31, 2022.

**Eligibility criteria:** Randomised controlled trials (RCTs) investigating the effects of BFFs on glucose, lipid, anthropometric, inflammatory and gut microbial parameters, in participants with obesity, MetS or T2DM.

**Data extraction and synthesis:** Two independent reviewers screened 6873 abstracts and extracted relevant data. Risk of bias (ROB) was assessed using the Cochrane Collaboration’s ROB2 tool. A qualitative, narrative synthesis was produced.

**Results:** The final review included 26 RCTs, with 31 reports published between 2001 and 2022. Significant (p<0.05) within-group and between-group changes in cardiometabolic outcome means were reported in 23 and 19 studies, respectively. Gut microbiota composition was assessed in four studies, with two finding significant between-group differences. No significant difference between groups of any measured outcomes was observed in five studies. There were 14 studies at low ROB; ten were of some concern; and two were at high ROB.

**Conclusion:** In 73% of included studies, BFF consumption by participants with obesity, MetS or T2DM led to significant between-group improvements in cardiometabolic outcomes, including fasting blood glucose, lipid profile, blood pressure, waist circumference, body fat percentage, and C-reactive protein. BFF consumption increased the abundance of beneficial gut bacteria, such as *Bifidobacterium* and LAB, whilst reducing potential pathogens like *Bacteroides*. To determine the clinical significance of BFFs as therapeutic dietary adjuncts, their safety, tolerability and affordability must be balanced with the limited power and magnitude of these preliminary findings.

**Ethics:** Ethical approval was not required as primary data was not collected.

**PROSPERO registration number:** CRD42018117766

**STRENGTHS AND LIMITATIONS OF THIS STUDY:**

- To the best of our knowledge, this is the first systematic review assessing RCTs of BFFs on metabolic, inflammatory, anthropometric and gut microbiota parameters in adults with obesity, T2DM, MetS or its components.
- Our search strategy adhered to the Cochrane review methodology and the PRISMA statement requirements.
- To ensure cultural inclusion and comprehensive up-to-date findings, our search started from inception to 31 August 2022, and had no language limits.
- ROB2, the most recent version of the Cochrane risk-of-bias tool, was used to assess the risk of bias in five domains covering the design, conduct and reporting of the included RCTs.
- Due to significant heterogeneity of BFF types, dosage, length of intervention and target populations, meta-analysis could not be conducted.

## INTRODUCTION

Metabolic syndrome (MetS) is an increasingly prevalent constellation of cardiometabolic derangements. Signified by central obesity, individuals with MetS suffer from atherogenic dyslipidaemia (raised triglycerides, lowered HDL-C), hypertension and impaired fasting glucose/ prediabetes.[1, 2] These interrelated risk factors are associated with an increased likelihood of developing type 2 diabetes mellitus (T2DM) and cardiovascular disease.[1, 3] Gut microbiota appears to be important in the pathogenesis of metabolic disorders;[4] adults with obesity and T2DM display markedly reduced gut microbiota diversity and low bacterial gene richness.[5–8] Diet regulates gut microbiota composition and function,[9, 10] and is a well-established modifiable factor in the management of MetS and T2DM.[11] Globally, MetS affects >25% of all adults.[12] Projections indicate that 1 billion adults will develop obesity by 2030,[13] with 700 million adults developing T2DM by 2045.[14] As such, dietary modulation of the gut microbiota may be a cost-effective approach to reducing the global health burden of obesity, MetS and T2DM.

Fermented foods are microbially-transformed foods traditionally consumed worldwide. Recent studies suggest that these foods, whether dairy or plant-based, deliver health benefits through transient integration of food-associated live microorganisms into gut commensal communities;[15] microbial enzymatic substrate transformation in the intestinal lumen;[16] the release of bioactive compounds including those with insulinotropic or immune-regulatory effects;[17] or the bacterial biosynthesis of vitamins.[18] Recent multi-omic approaches have found that fermented foods may be an important source of commensal lactic acid bacteria (LAB) in the gut microbiome,[19] and that consumption of large amounts of fermented foods leads to subtle, persistent differences in human gut microbiota composition and faecal metabolome.[20] In a recent study, diets which were high in fermented food increased microbiota diversity and reduced inflammatory markers compared to plant-based diets with twice the fibre and no fermented foods.[21] Abundant clinical studies investigating the effect of fermented dairy products on cardiometabolic health have been systematically reviewed and meta-analysed,[18] with mixed results. Overall, fermented milk consumption is associated with a reduced risk of T2DM and may assist with weight maintenance and obesity.[22–24] However, less well studied are botanical fermented foods (BFFs), which may be even more effective at exerting health effects than their dairy-based counterparts.[25]

BFFs are globally consumed plant-based foods and beverages,[16, 26] produced through the fermentation of vegetables, fruit, cereals, nuts and pulses. Traditional BFFs include kimchi, sauerkraut, water kefir and tempeh. Compared to dairy-based fermented foods, BFFs may contain more diverse microbial communities, with more microbial genes (gastrointestinal survival, gut colonisation, immune modulation) associated with potential health benefits to the host.[27, 28] BFFs are also lower in fat; cholesterol free; and contain higher levels of polyphenols, other antioxidants and microbiota-associated carbohydrates, including dietary fibre.[29, 30] They are also better tolerated and more palatable for many consumers,[31] especially as 75% of the world’s population is lactose intolerant.[32] For these reasons, BFFs are potentially more suitable than fermented dairy products for the prevention and management of obesity, MetS and T2DM.

Recent critical reviews of fermented foods have stated that more human studies are required to establish the role of BFFs as interventions for noncommunicable diseases.[18,33,34] However, as these reviews did not use systematic review methodology, many existing clinical trials of BFFs were not identified. On the other hand, we found several up-to-date systematic reviews/ meta-analyses of randomised controlled trials (RCTs) investigating the impact of red yeast rice/ monacolin/ xuezhikang [35–39] or vinegar [40–45] on metabolic parameters; these RCTs were thus excluded during abstract screening to prevent the waste of resources.[46] We have undertaken a systematic review of RCTs administering other BFFs to participants with T2DM, MetS, obesity, or other MetS components. Our review asks: “Does the consumption of BFFs have any impact on obesity, T2DM or MetS outcomes, including the gut microbiota?”

## METHODS

### Search strategy

We performed a qualitative systematic review of randomised controlled trials (RCTs) conducted in adults with type 2 diabetes mellitus (T2DM) or metabolic syndrome (MetS) components or any combination of these components (Online Supplemental material 1). This systematic review was registered at the International Prospective Register of Systematic Reviews (PROSPERO) website on 18 December 2018 (CRD42018117766). The detailed methods of this review were reported in a protocol paper.[47] Embase, MEDLINE, Cochrane CENTRAL and Google Scholar (first 200 relevancy ranked results) were searched from inception to 30 August 2022. Our search strategy (Online Supplemental material 2) combined subject heading terms and text words for BFF (e.g. fermented food, fermentation), and MetS or T2DM (e.g. MetS, obesity, hypertension, blood pressure, diabetes, prediabetes, hyperlipidaemia, microbiota, dysbiosis, inflammation). To retrieve RCTs, the Cochrane Highly Sensitive Search Strategy for MEDLINE [48] was used. No language limits were applied. Reference lists in identified articles were searched with Scopus. We also searched grey literature via trial registries and conference papers. When a study had unreported data, authors were contacted for further information.

### Selection criteria and data management

Reviewers MC and NL independently selected and extracted data from eligible publications. Selection was based on the PICOS criteria,[49] as shown in Table 1. The following data was extracted: first author’s name, publication year and study location; study design; BFF and comparator type, dosage and duration; subject characteristics (sample size, population, age, gender, condition); metabolic and gut microbiota parameters (mean ± SD/SE) in each group before and after the intervention. Study populations described in more than one published article were counted as single studies, with relevant data extracted from all articles. Any discrepancies were resolved through discussion. Authors of trials were contacted for clarification when necessary. All processes and data were recorded using Covidence software (www.covidence.org). The PRISMA literature search [50] results are represented in Figure 1.

**Table 1.**
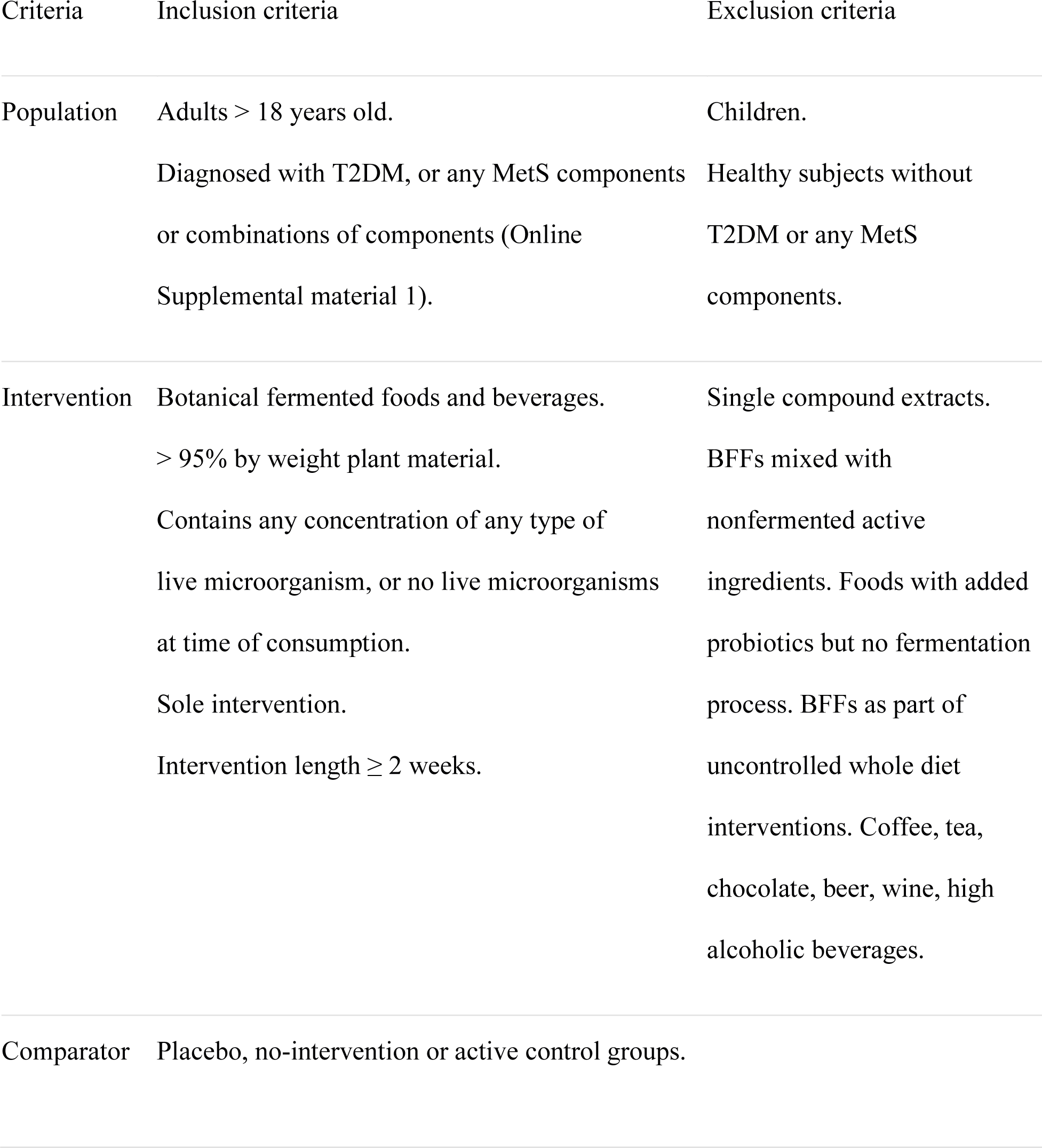
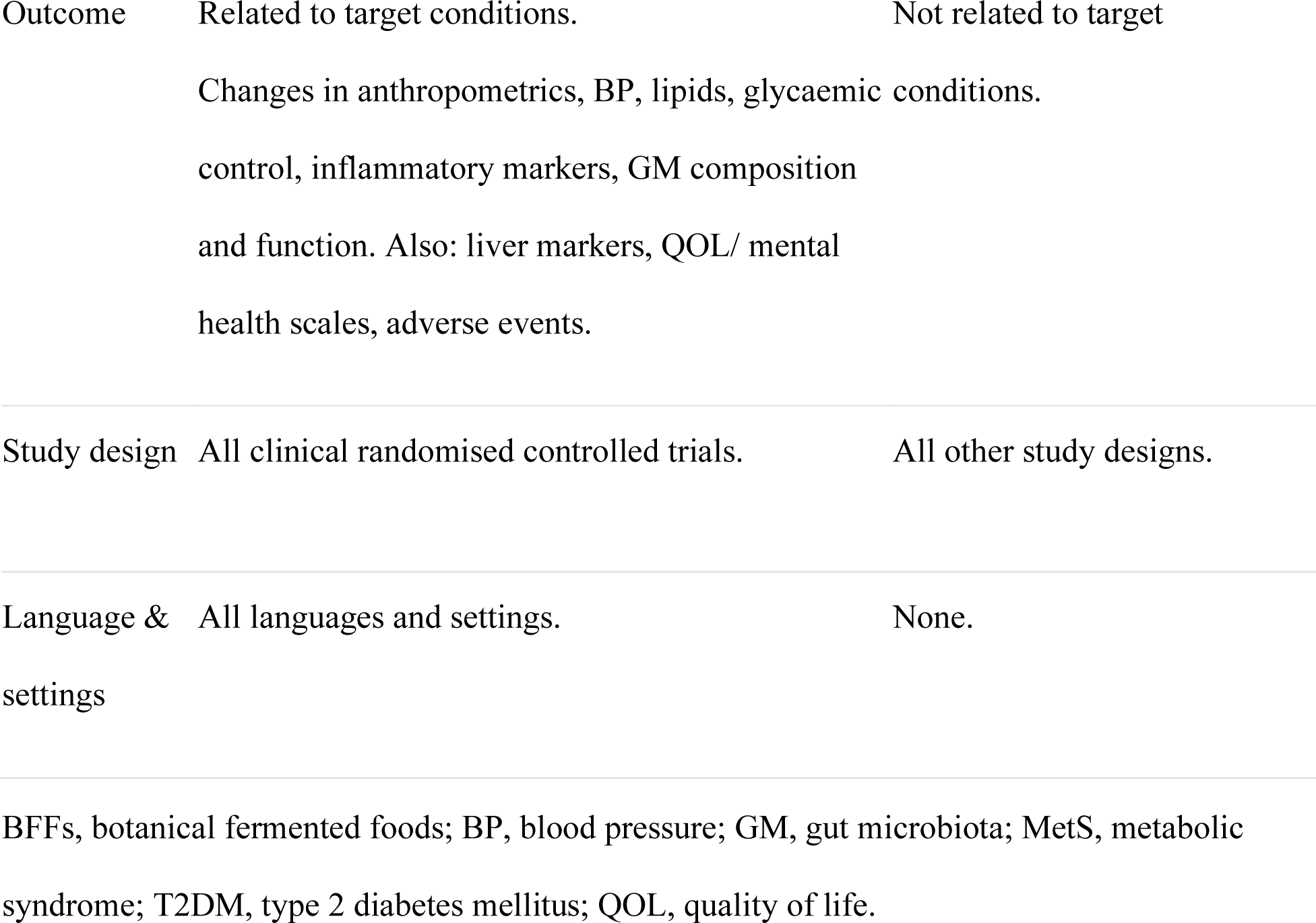
PICOS (population, intervention, comparison, outcome, study design) criteria for inclusion of studies.

**Figure 1.**
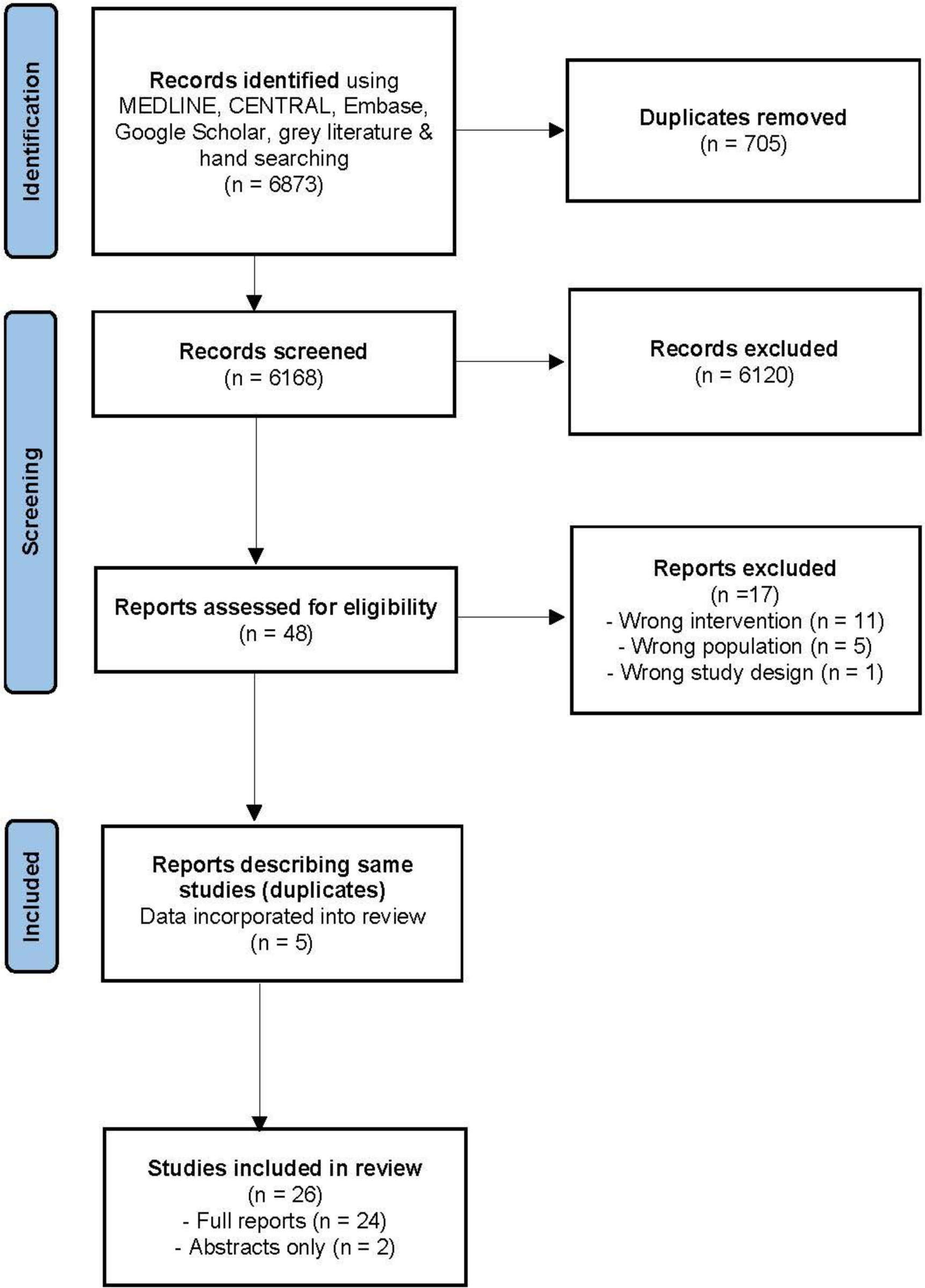
PRISMA flow diagram of literature search.

Primary outcomes were changes in any T2DM and MetS clinical parameters. Anthropometric measures were body weight (BW), Body Mass Index (BMI), waist circumference (WC), waist-hip ratio (WHR), body fat percentage (BFP), total fat area (TFA), visceral fat area (VFA), subcutaneous fat area (SFA), systolic and diastolic blood pressure (SBP, DBP). Lipid profile consisted of total cholesterol (TC), triglycerides (TG), high-density lipoprotein (HDL-C) and low-density lipoprotein cholesterol (LDL-C), and free fatty acids (FFA). Glucose metabolism was measured via fasting blood glucose and insulin (FBG, FBI), 2-hour postprandial glucose and insulin (2hPPG, 2hPPI), haemoglobin A1c (HbA1c), Homeostatic Model Assessment for Insulin Resistance (HOMA-IR) and C-peptide (C-pep). Secondary outcomes were shifts in: inflammatory markers interleukin-6 (IL-6), interleukin-10 (IL-10), tumour necrosis factor alpha (TNF-*α*), C-reactive protein (CRP) and high sensitivity-CRP (hs-CRP); obesity markers apolipoproteins A1 (ApoA1) and B (ApoB), and ApoB/A1; gut microbiota composition; and liver function tests.

### Risk of bias assessment

All included studies were independently qualified by MC and NL using the Cochrane Collaboration’s Risk of Bias 2 (ROB2) tool [51] within Covidence. Each study was assessed as having a low risk of bias, some concerns, or a high risk of bias.

### Data synthesis strategy

We deemed a meta-analysis inappropriate due to between-study clinical heterogeneity in participant groups, diagnostic cut-off points, interventions, comparators and outcomes measured. We presented a narrative synthesis, organised according to fermented food/ microorganism types: 1) lactofermented foods, produced through the fermentative action of LAB, such as kimchi and ash kardeh; 2) jangs, such as kochujang, doenjang and chungkookjang, produced by fermenting soybeans with *Bacillus* and *Aspergillus* spp.; 3) tempeh, produced by fermenting soybeans with *Rhizopus* spp.; 4) *Aspergillus oryzae*-fermented products, such as shiokoji, amazake and miso; and 5) others. For more detail on the production of these fermented foods, please see Online Supplemental material 3. Results were then described according to relevant biochemical and clinical parameters (lipid, glucose, anthropometric, inflammatory and gut microbiota parameters), as well as populations studied (overweight/obese; impaired fasting glucose/ prediabetic/ T2DM; hypertension; hyperlipidaemia; and MetS/ cardiovascular risk factors).

## RESULTS

Our search yielded a total of 6873 citations. Following removal of duplicates, abstract screening and full-text review, 31 published reports of 26 RCTs met our inclusion criteria. Relevant data was extracted from all reports and combined for the same study when duplicates were present.

## Assessment of risk of bias

Using the Revised Cochrane risk-of-bias tool (ROB 2), we assessed all included studies for risk of bias. The results of judgments by two investigators, according to each of the tool’s five domains, are summarised in Figures 2 and 3, produced using the Robvis tool.[52] Bias due to randomisation was high risk in 8% of studies, of some concern in 31%, with 61% at low risk. The studies showed a low risk of bias in the following domains: deviations from intended interventions (100%), missing outcome data (92%), outcome measurement (100%), and selection of reported result (92%). Overall risk of bias was low in 54% of studies, of some concern in 38% and high in 8%.

**Figure 2.**
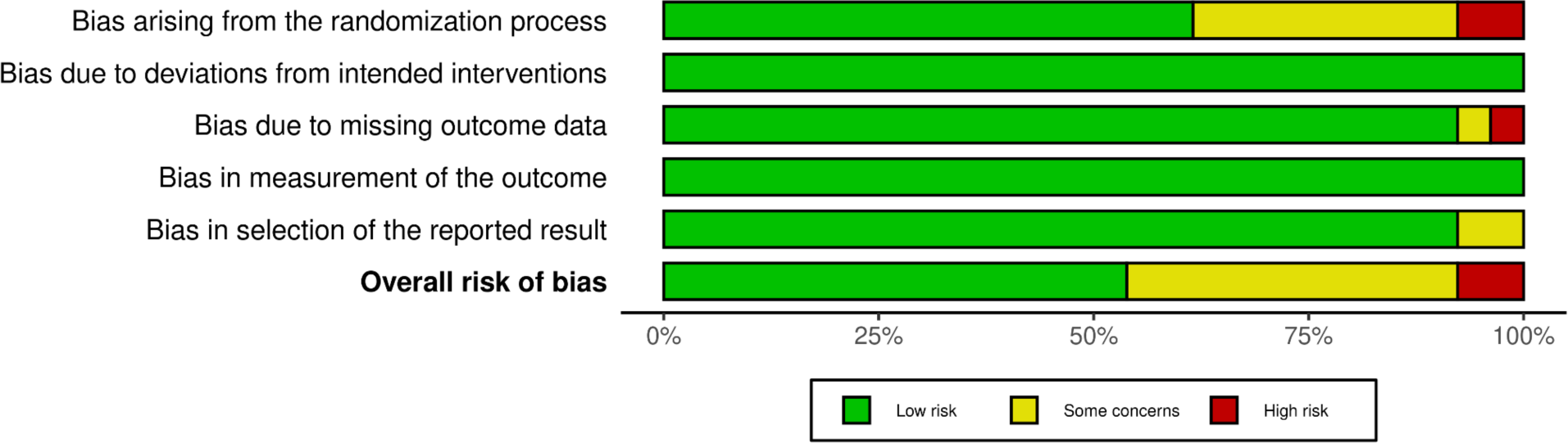
Risk of bias summary: domain-level judgements for each individual study.

**Figure 3.**
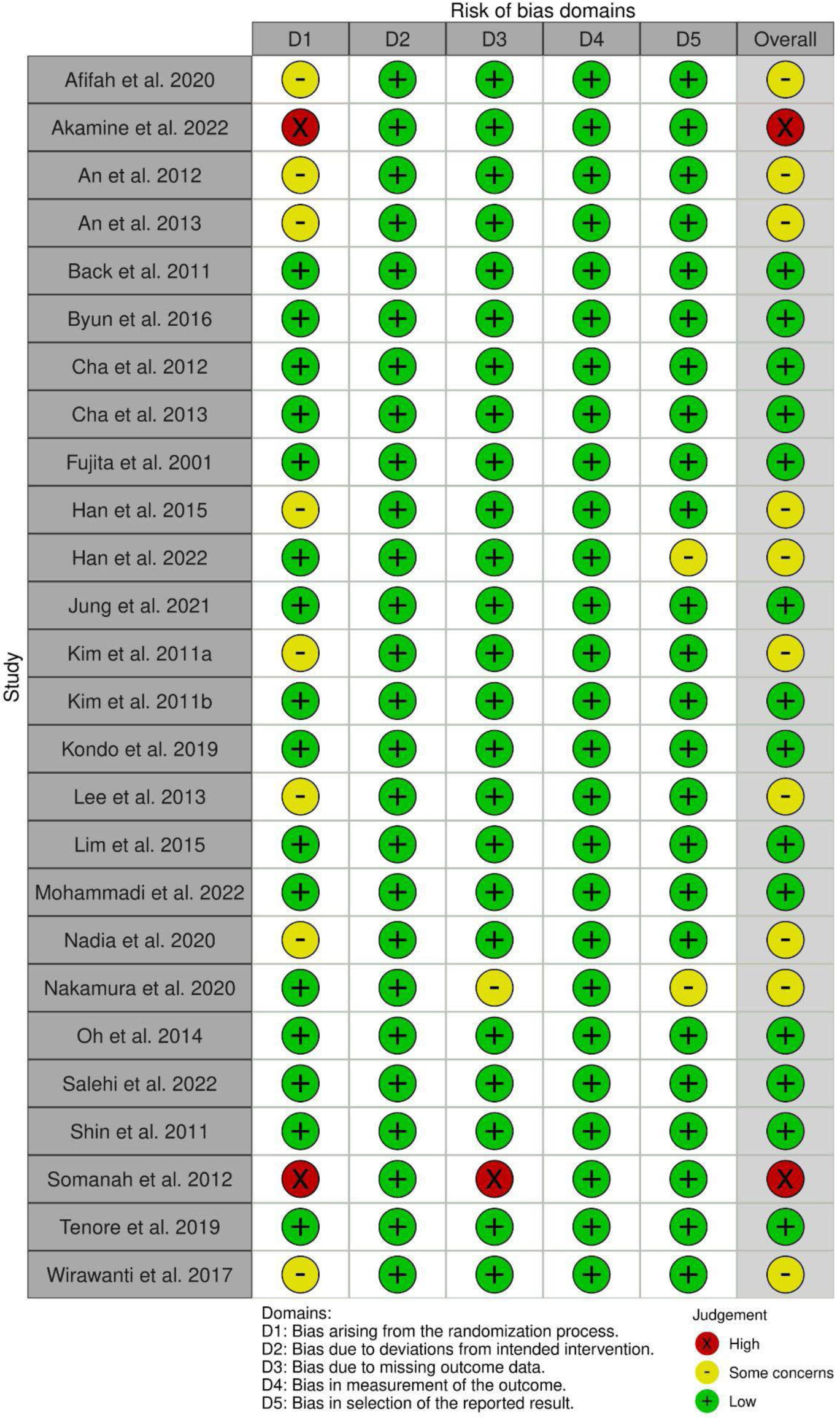
Risk of bias graph: weighted distribution of risk-of-bias judgements within each bias domain across all included studies.

Risk of bias during the randomisation process was low in all but ten studies: eight were of some concern,[53–60] and two were at high risk.[61, 62] These ten studies did not describe randomisation or allocation concealment methods in enough detail. All studies had almost equal baseline numbers in each intervention group, with the exception of one study [61] which did not use block randomisation and had substantial differences in group size allocation (intervention group n=49; control n=78). Akamine et al. [62] reported significant baseline imbalances in FBG levels (brown rice amazake 5.6 ± 0.2 vs. white rice amazake control 6.3 ± 0.2 mmol/L; p=0.018); the number of subjects with T2DM (brown rice amazake n=3 vs. control n=10; p=0.017); and, gut microbiota composition at genus and species levels (*Bacteroides intestinalis*, *Faecalibacterium spp. DJF VR20*, *Sutterella wadsworthensis*, *Parabacteroides distasonis*, *Alistipes onderdonkii* were significantly higher in the brown rice amazake group; p<0.05). Kim et al.,[53] Somanah et al. [61] and An et al. [55] did not provide sufficient information on baseline imbalances between groups, as well as An et al. [54] and Lee et al.,[56] which were abstracts only. Han et al. [57] stated that “CRP and DBP are significantly different between groups”. All other studies showed no baseline imbalances between groups.

No studies reported toxicities or serious adverse events, likely due to the interventions being foods or food based. As such, although there were 11 open-label studies,[53–63] no studies were at risk of bias due to deviation from intended interventions.

No dropouts were reported in four studies.[53,60,64,65] Akamine et al. [62] also reported no dropouts, but only analysed 88% of participants for plasma SCFA, and 85% of participants for faecal microbiota composition, due to missing or uncollected samples. Analysis of over 90*%* of randomised participants (91 to 96%) was conducted in ten studies;[55,57–59,63,66–70] all of these studies used per-protocol analysis. Between 80 to 90% of participant data was analysed in five studies,[71–75] with only Cha et al. [72] and Oh et al. [73] using intention-to-treat analysis for missing data. Tenore et al.,[76] with only 69% of participants completing the study, used a “negative binomial, generalised linear mixed effects model with a per-protocol set” to account for missing data. The remaining studies included data analysis for 79%,[61] 69%,[77] and 61% [78] of participants. Somanah et al.’s [61] study was high risk in the domain of missing outcome data as there were large numbers of dropouts, combined with unexplained inconsistencies in the number of participants analysed for each outcome. Nakamura et al. [78] provided primary analysis data (shiokoji n=23; placebo n=24) as per their original inclusion criteria, finding no significant differences between groups in glucose parameters. On completion of the study, investigators excluded participants who had significant fluctuations in physical activity or alcohol intake during the trial (seven from each group), and/ or those with high insulin resistance (HOMA-IR > 5.0) (two from placebo group HOMA-IR >5.0; one from placebo group with both factors). Secondary post-hoc analysis using the same pre-specified statistical methods was performed, resulting in a significant finding. Although both analysis sets were provided, reasons were documented and limitations stated, it is difficult to ascertain if these decisions were made to elicit a positive result. As such, this study was assessed as having some concerns due to missing outcome data and selective reporting. Han et al. [70] was found to be of some concern regarding selective non-reporting, as it did not present any between-group analysis data; we were unable to ascertain, despite inquiries, whether analysis was performed, or if no significant differences were found and therefore unreported. In all other studies with dropouts, the authors documented the reasons for missing outcome data, which were all unrelated to the outcomes (e.g. noncompliance, personal reasons). None of the other studies used selective reporting, and all studies had a low risk of bias in measurement of outcomes.

Overall, we judged 14 of the included RCTs to have a low risk of bias.[63–69,71–77] Eight of the ten studies assessed to be of some concern [53–60] would likely be upgraded to low risk if more information regarding randomisation and allocation concealment was provided. Han et al. [70] likely engaged in selective non-reporting of between-group analysis; more information from authors may change the level of risk assigned. Nakamura et al. [78] would be deemed low risk if the primary analysis is used. Two studies were at high risk of bias: Somanah et al. [61] had a high risk of bias in the domains of randomisation and incomplete outcome data; Akamine et al. [62] had a high risk of selection bias as they did not conceal allocation, used “pseudo-randomisation”, and had a significant baseline imbalance in FBG levels, with unequal group numbers (19 vs. 21). In future studies, to reduce the risk of bias, intention-to-treat analysis should be utilised to account for missing data, and more details regarding randomisation, allocation concealment and baseline imbalances need to be included.

### Study characteristics

Table 2 summarises the characteristics of included RCTs. RCTs by An et al. [54] and Lee et al. [56] were only described in abstracts. There were seven crossover studies,[53–56,69,75,77] with the rest being of parallel design; 15 were double-blinded or more,[64–78] whilst 11 were open-label studies. Women only were recruited in three studies;[57,59,60] all other studies included both genders. Participants were between the ages of 19 [77] and 65 [66]. The sample sizes were between 16 [56] and 127 [61], with intervention periods ranging from 2 [59] to 14 [61] weeks, with variable BFF doses, and no statement of microbial counts, except in two studies.[53, 76] A variety of BFFs fermented with different microorganisms, from LAB, *Bacillus*, *Aspergillus* and/ or *Rhizopus* spp., were used as interventions. Comparators included placebos, no intervention and active controls. Some studies provided meals to both groups, and dietary intake data was collected and analysed across all studies. The populations investigated in the studies originated from South Korea (n=14), Japan (n=4), Indonesia (n=3), Iran (n=2), USA (n=1), Italy (n=1) and Mauritius (n=1).

**Table 2.**
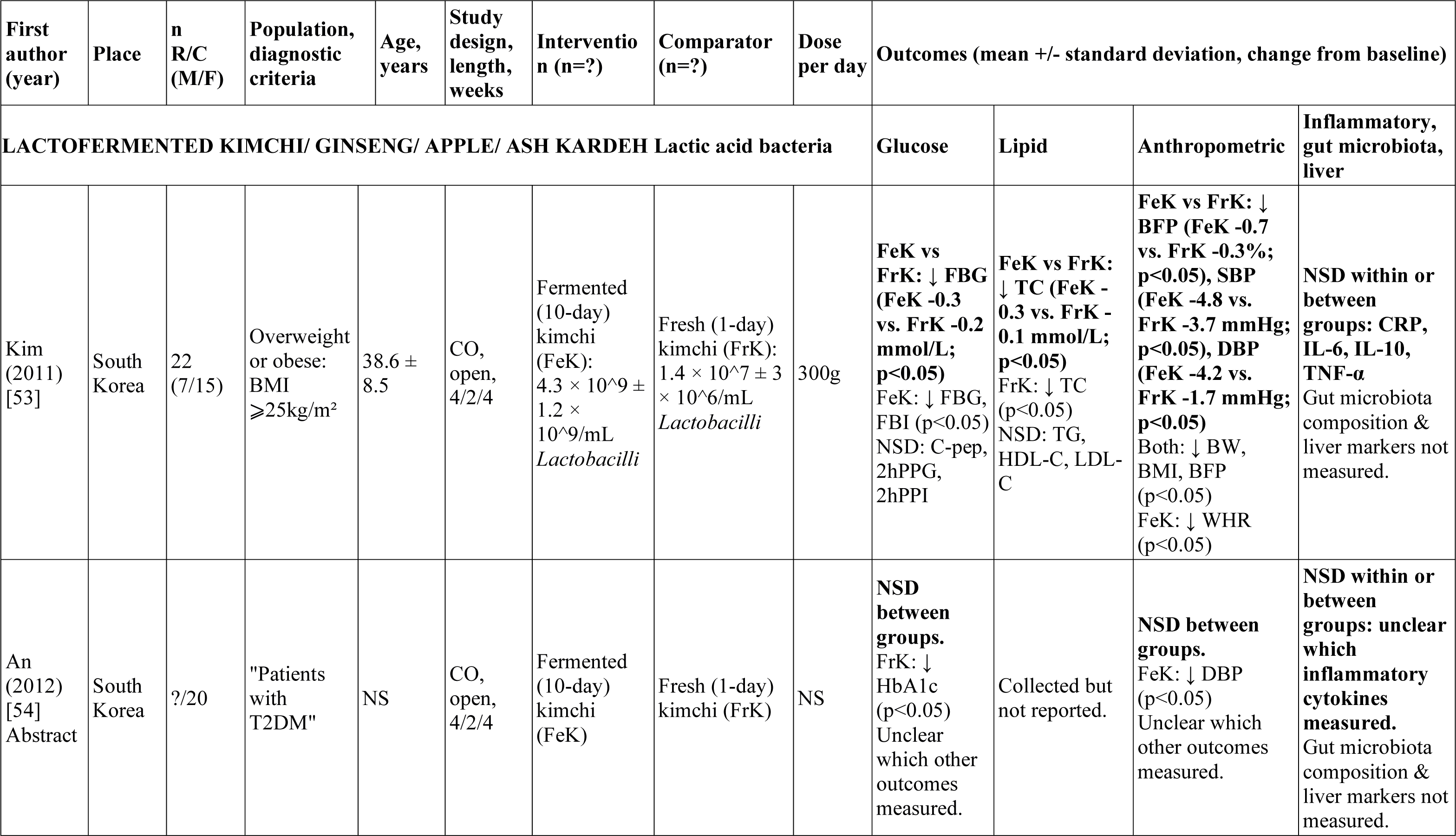
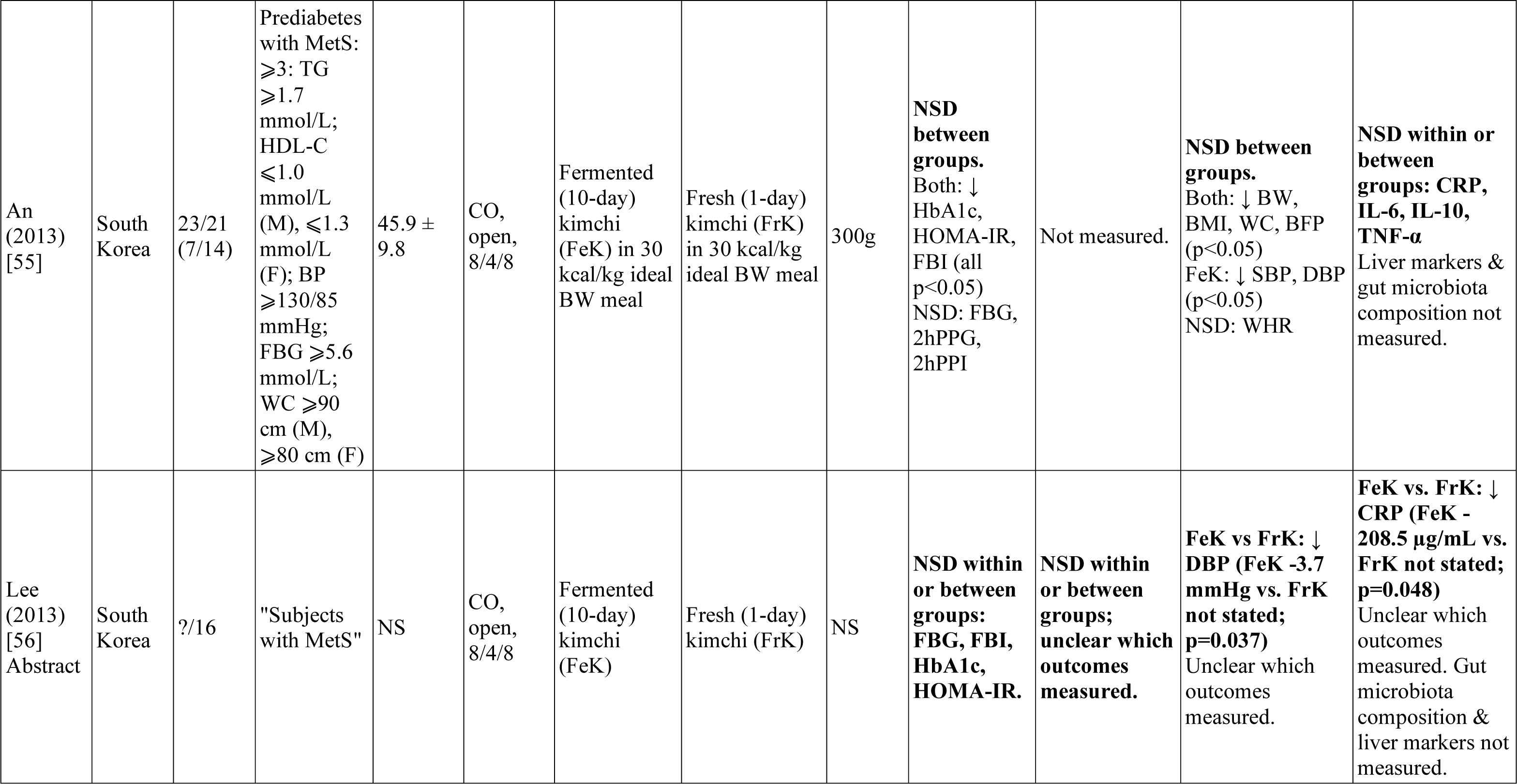
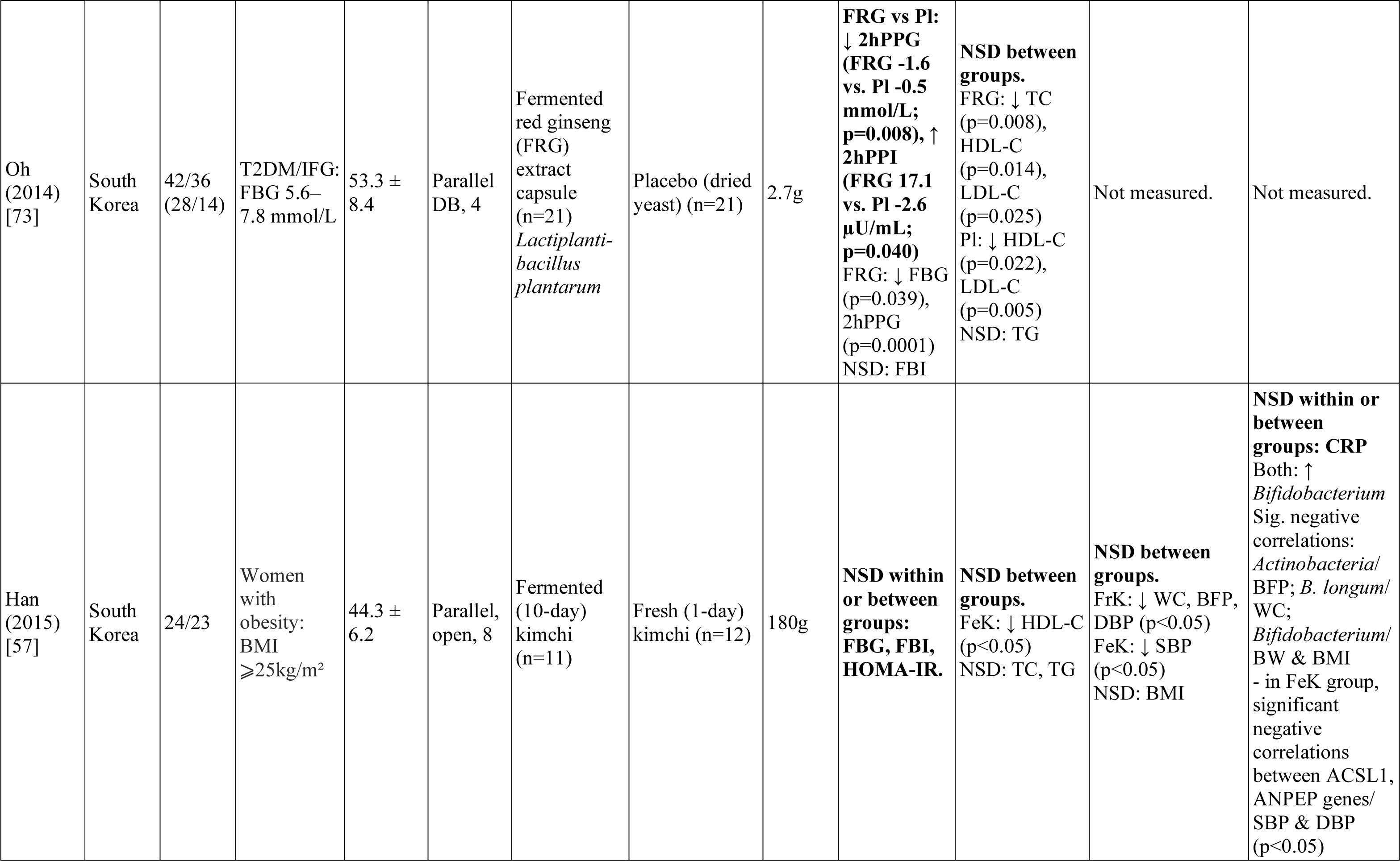
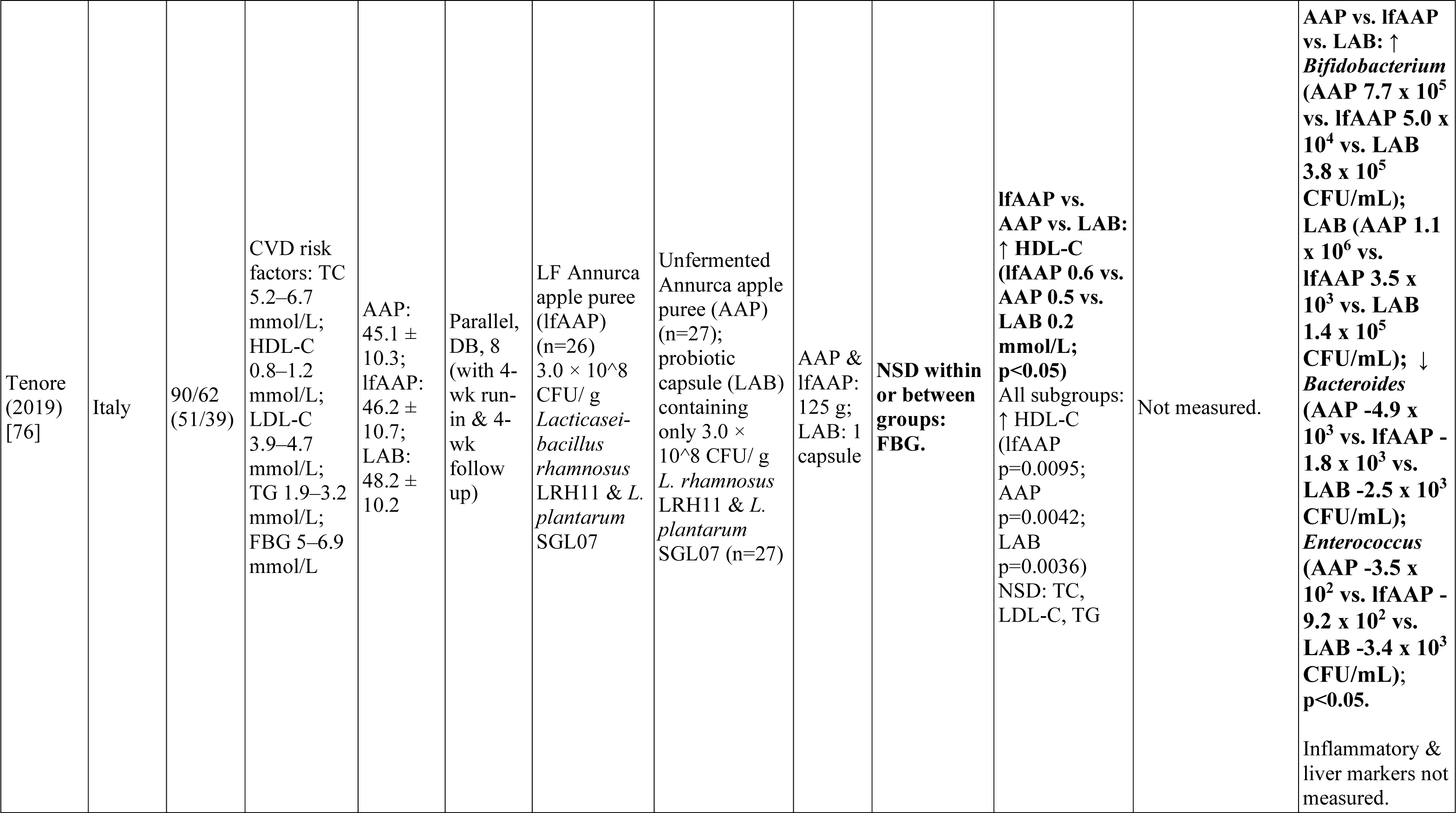
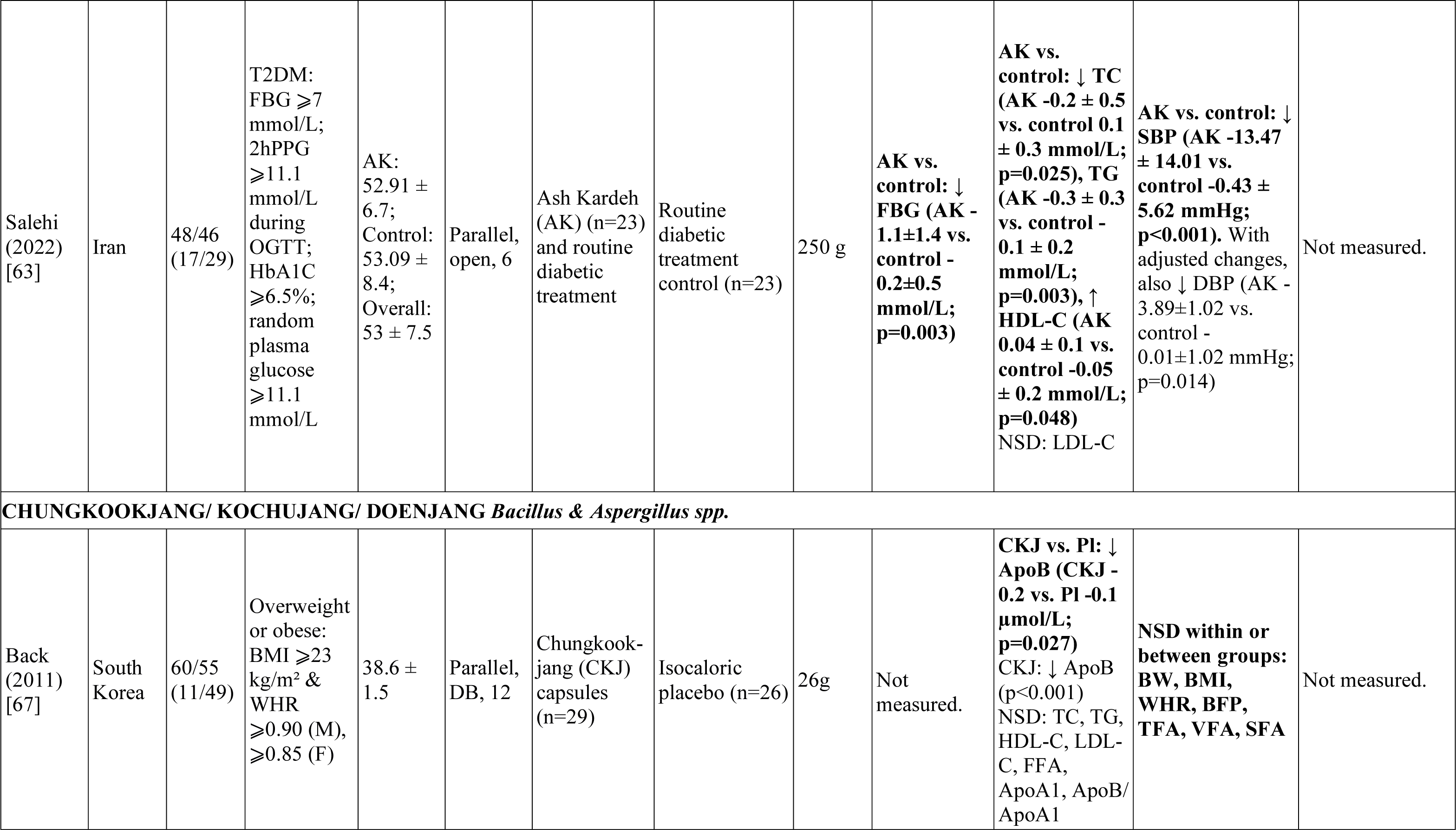
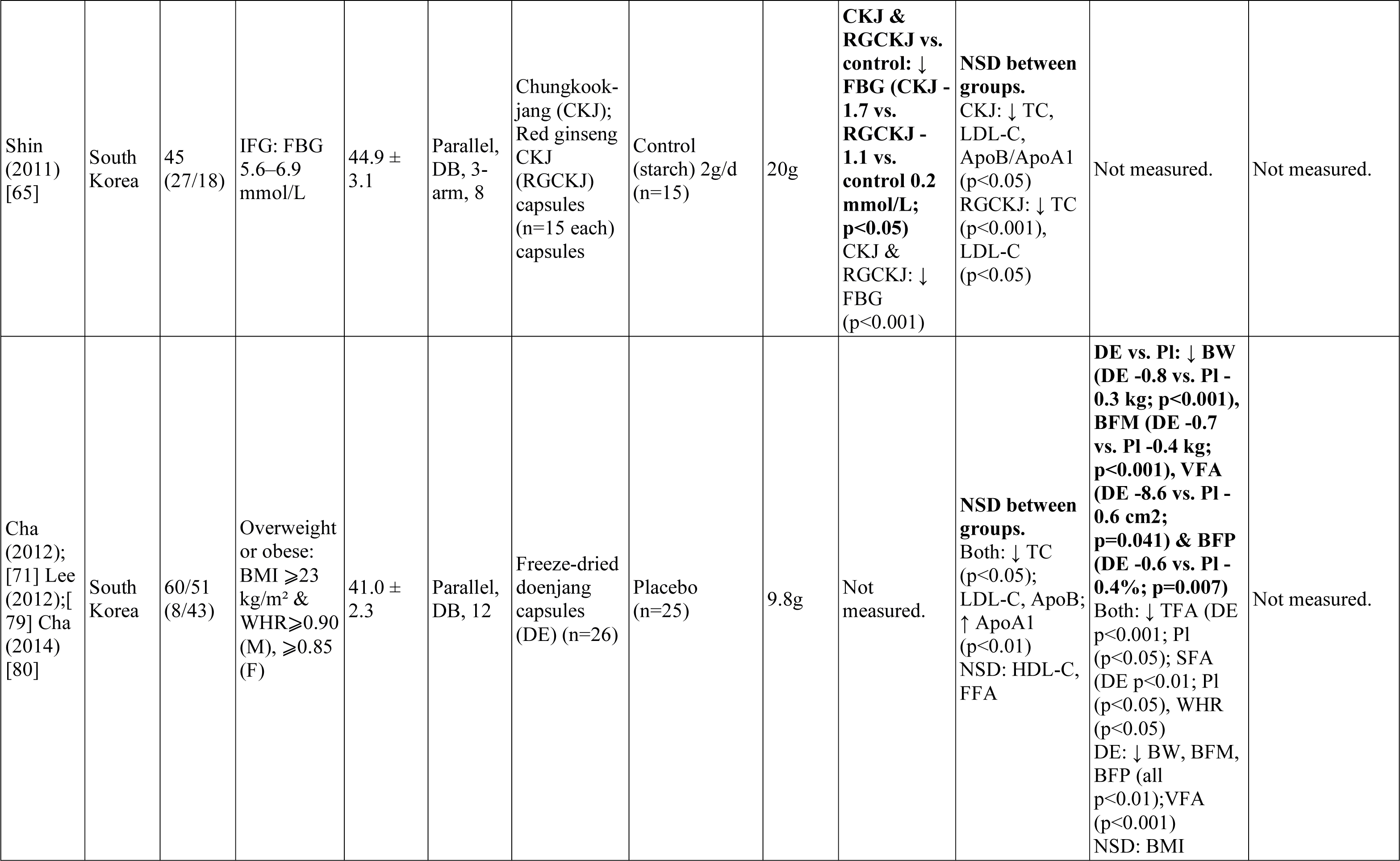
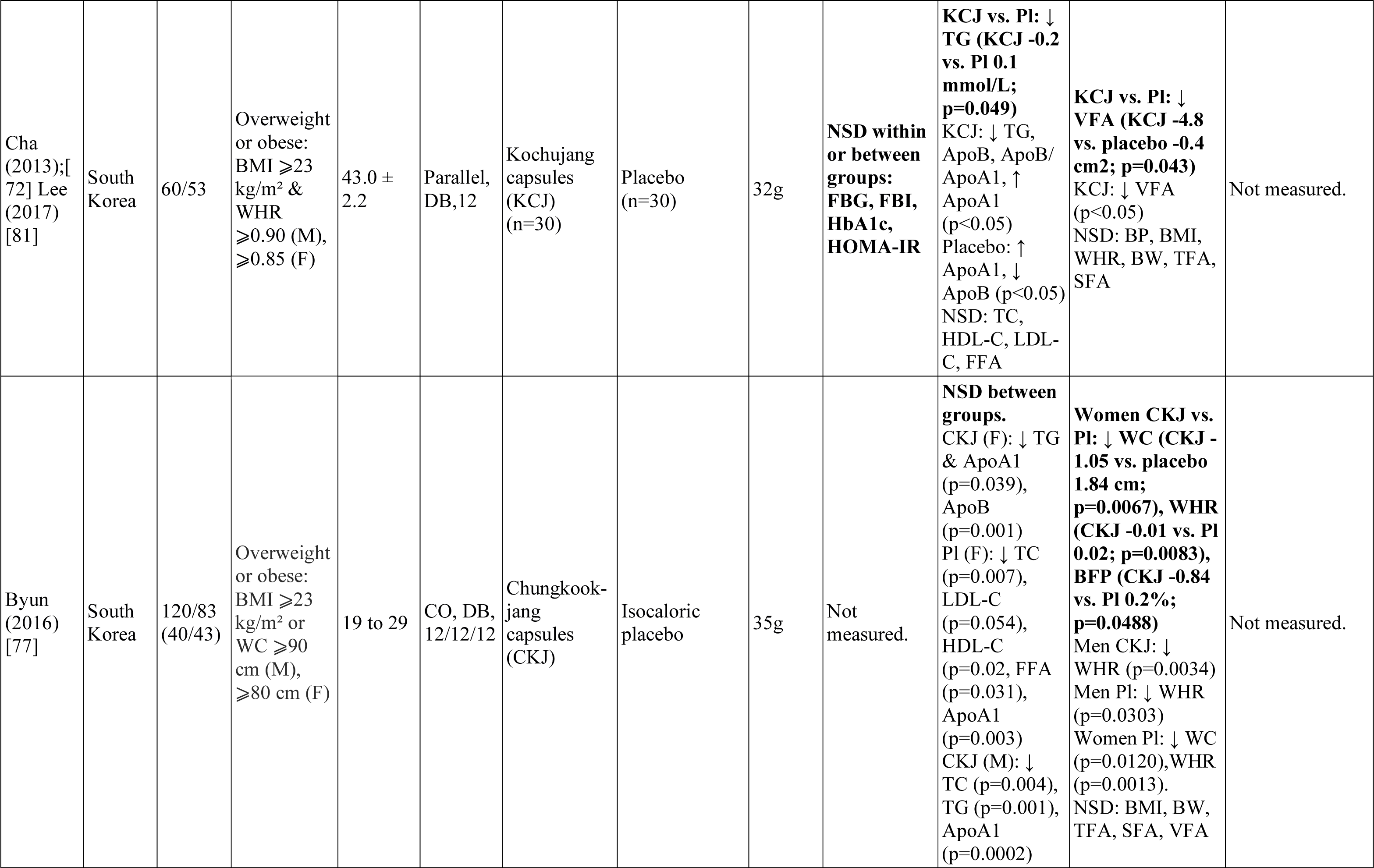
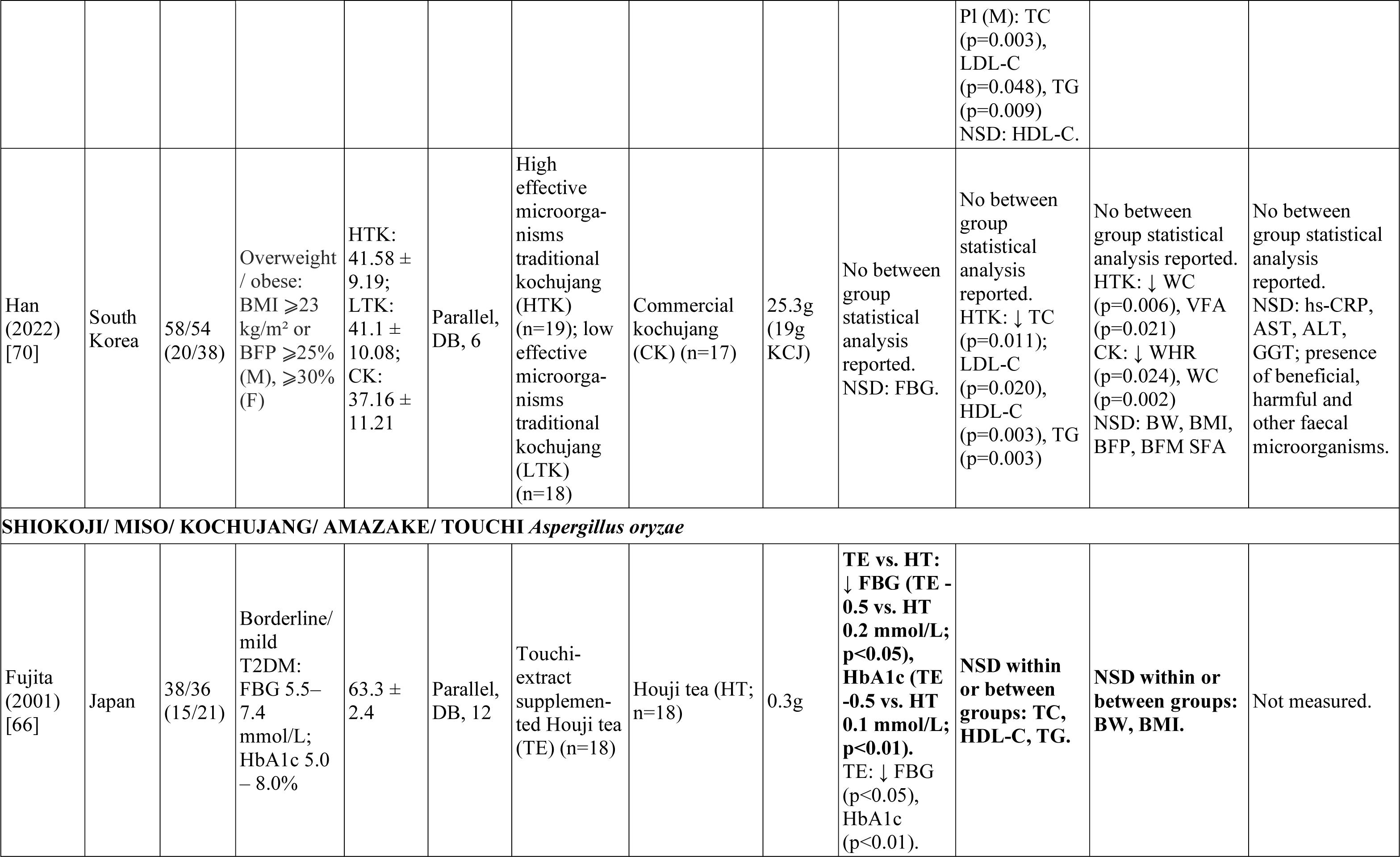
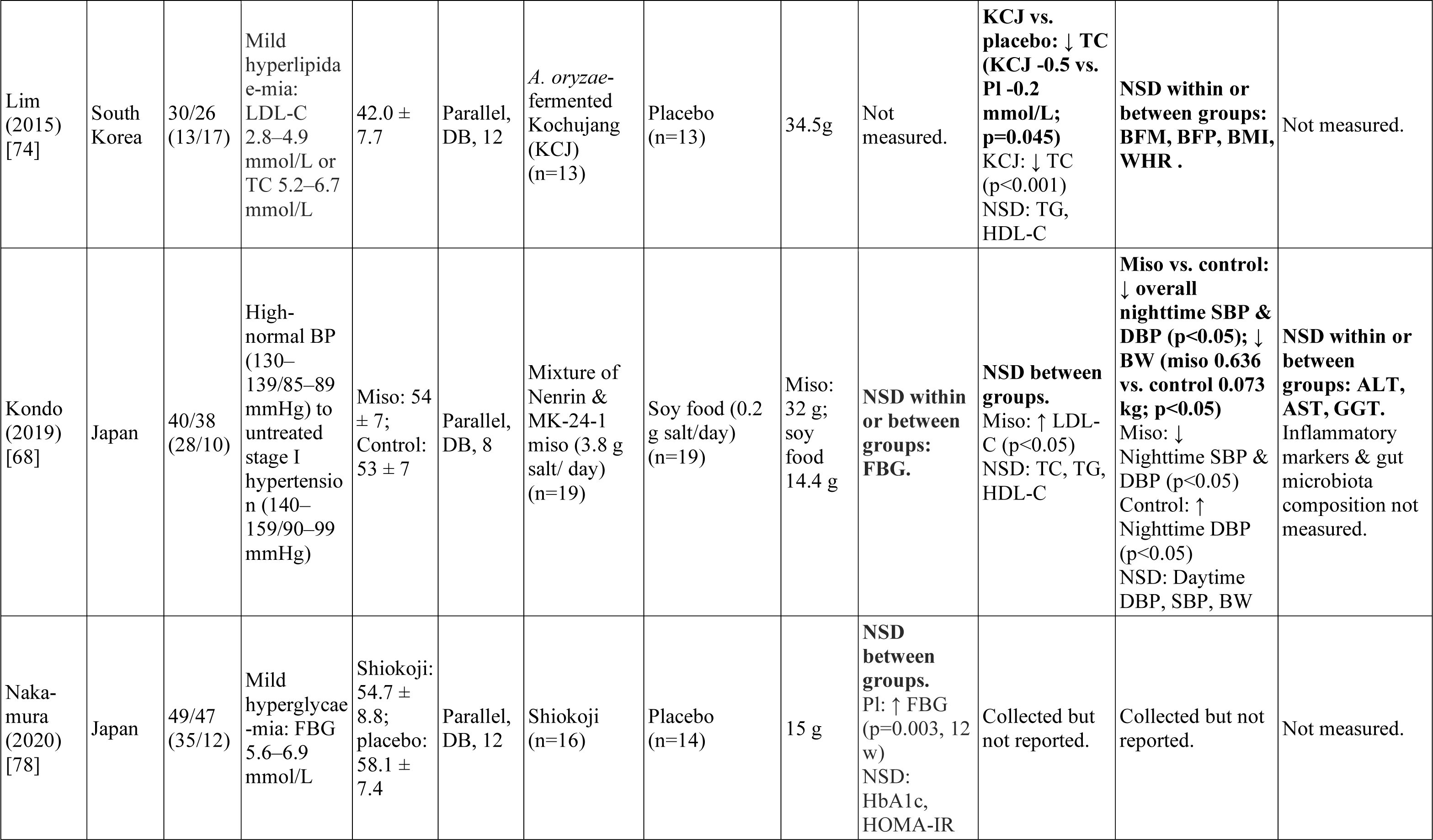
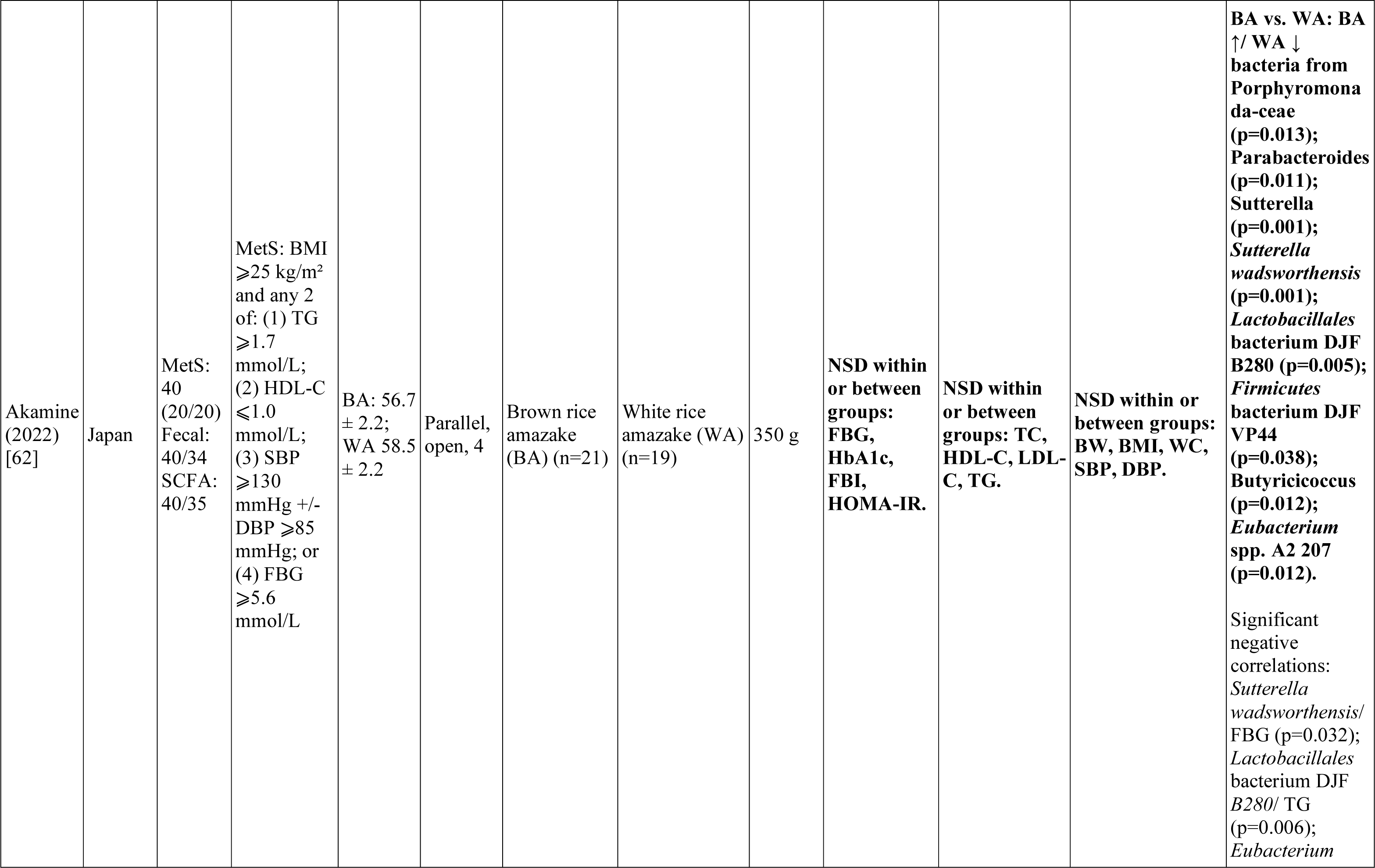
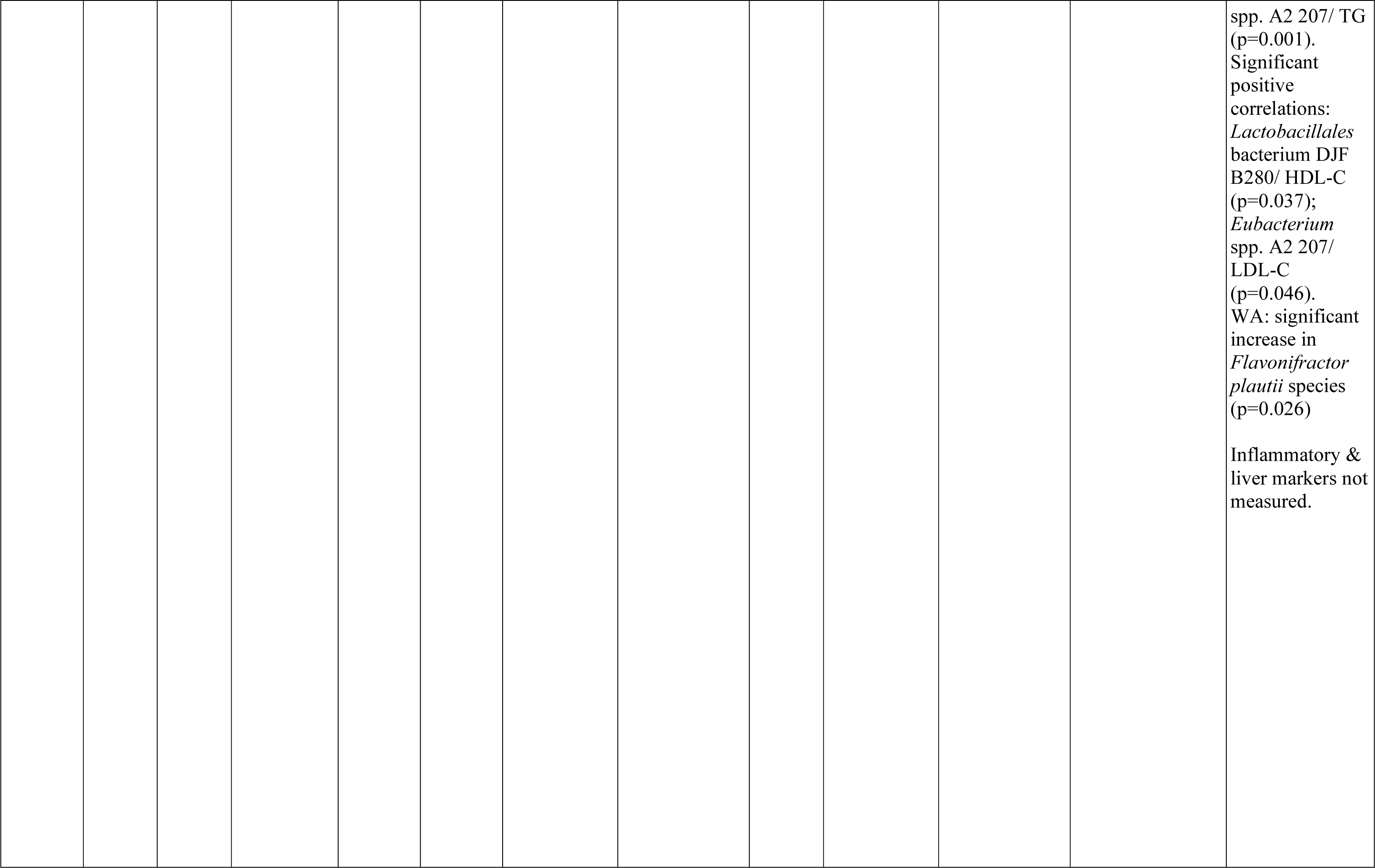
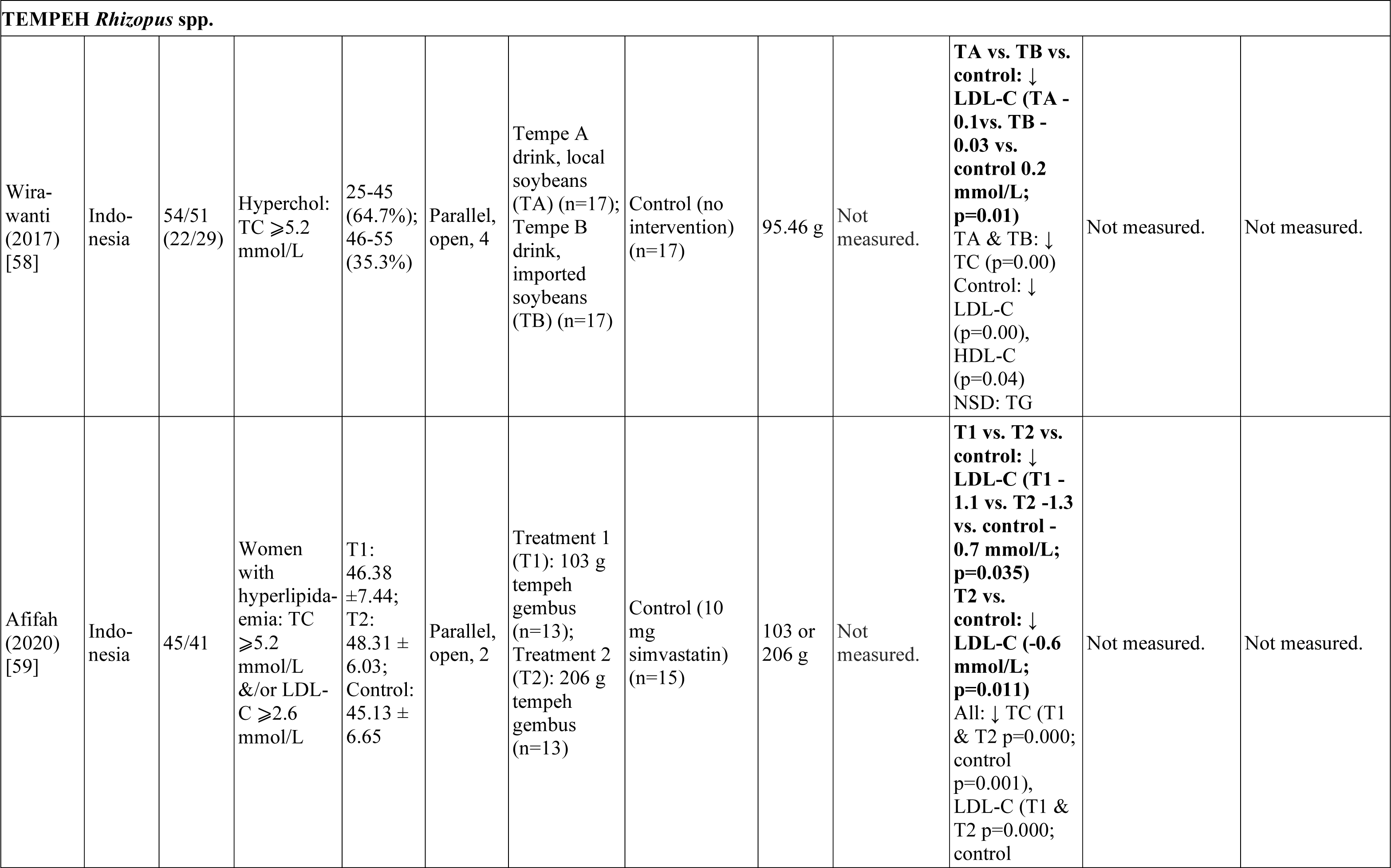
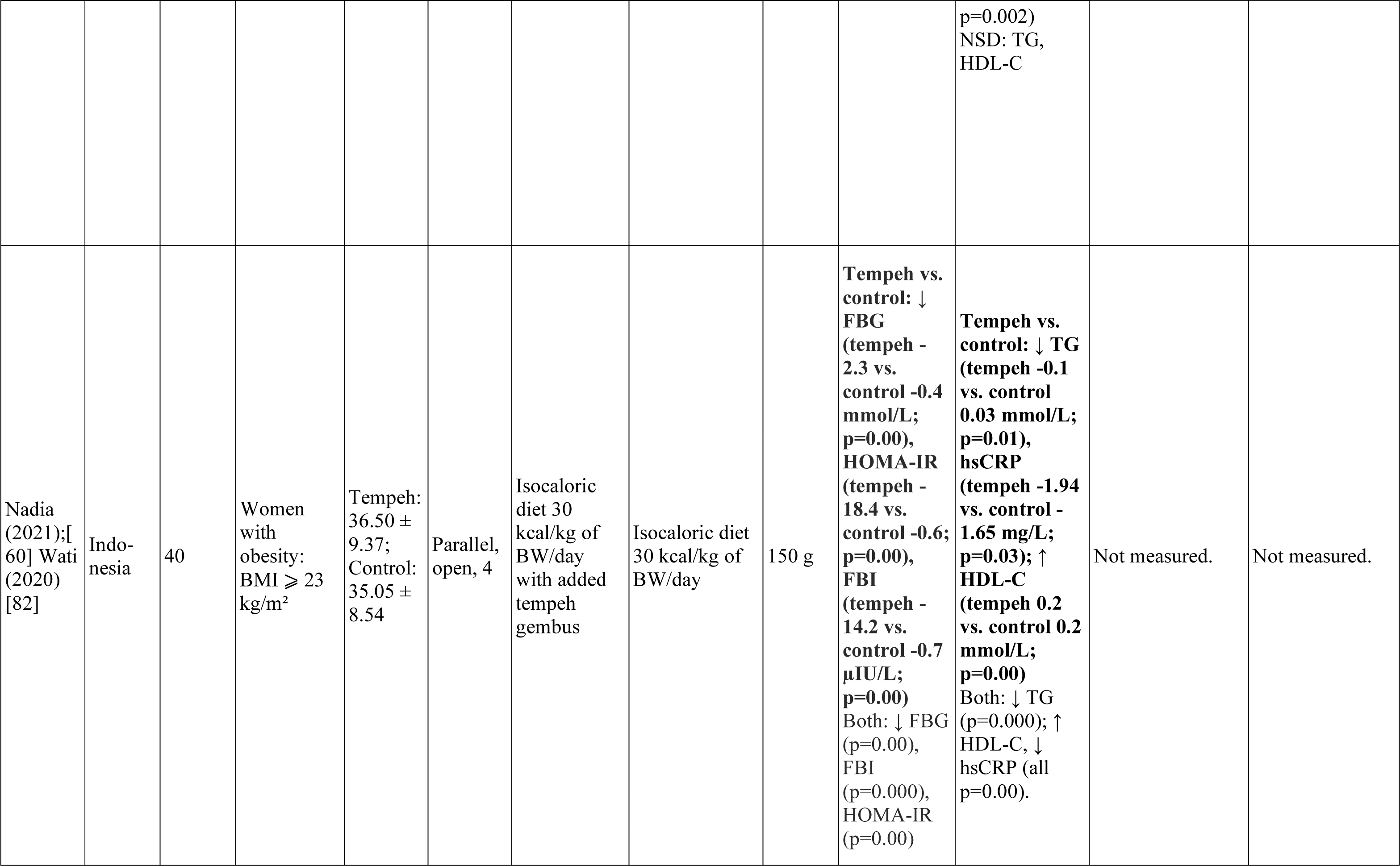
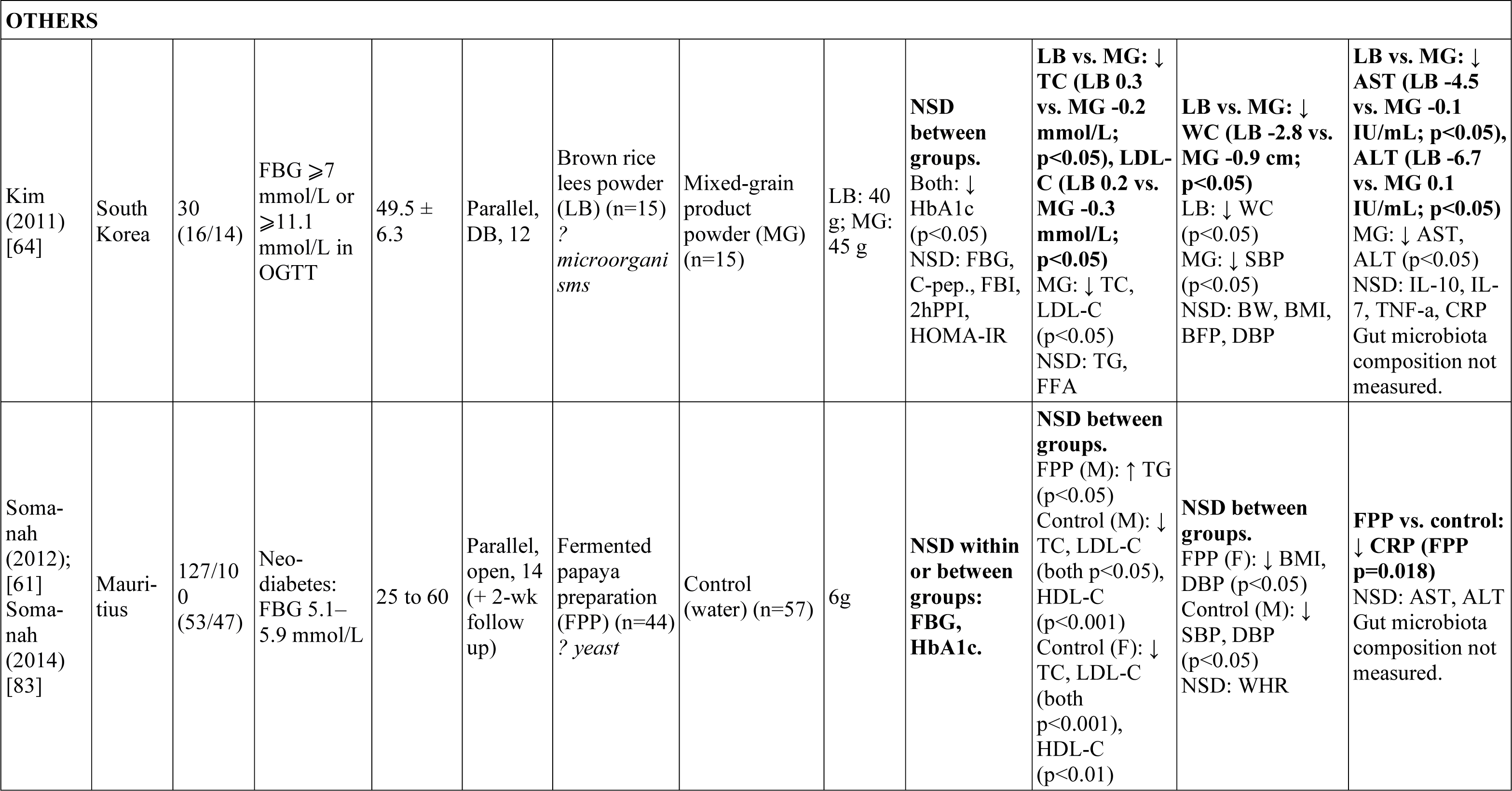
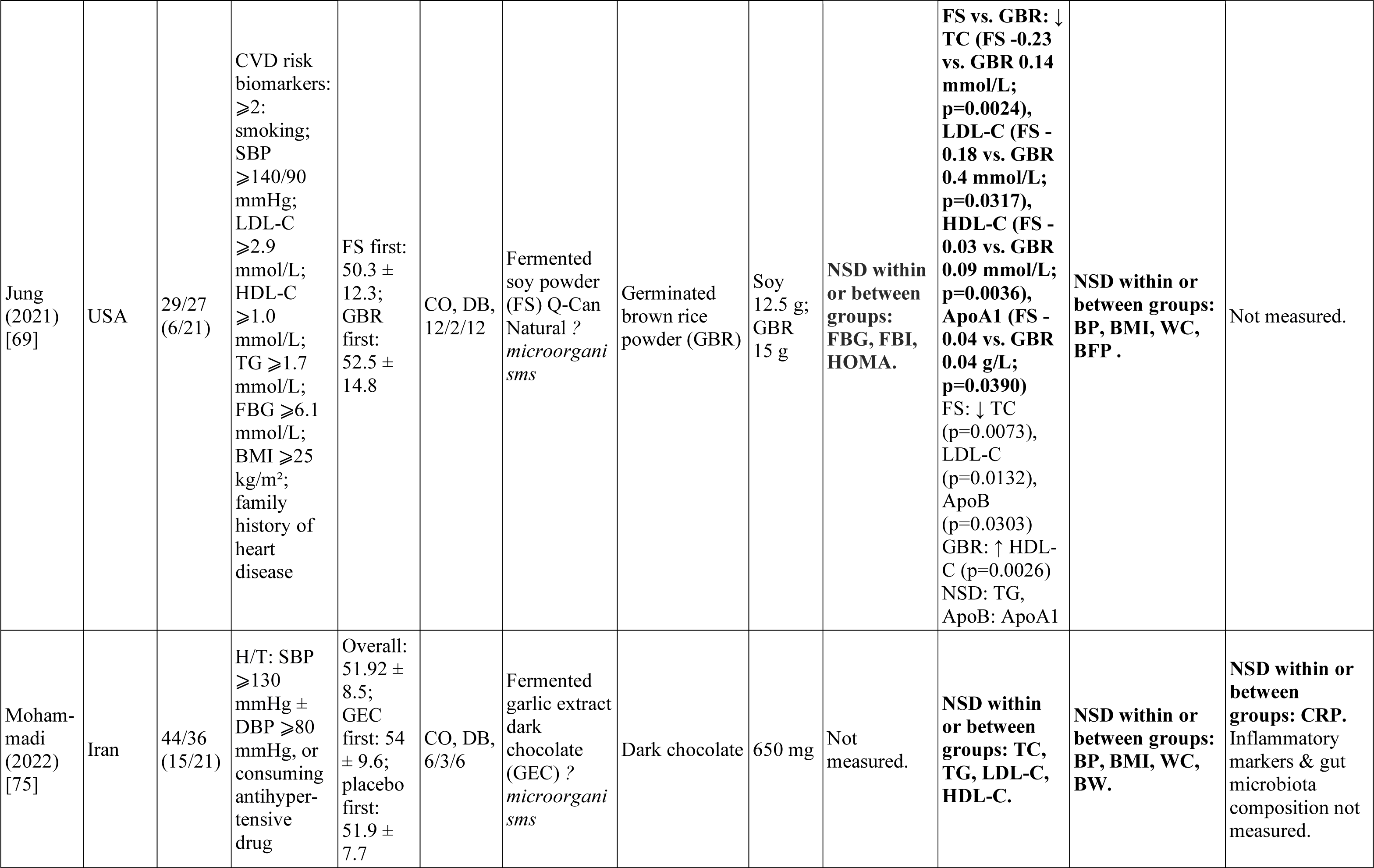
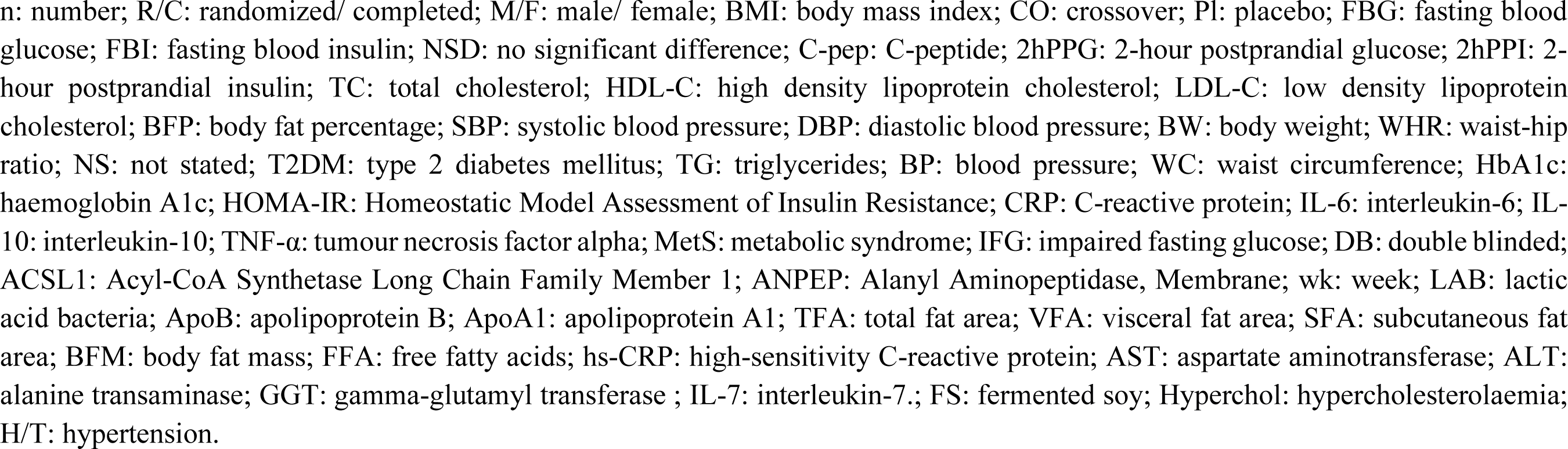
Characteristics of included randomised controlled trials.

The studies measured some combination of the following relevant outcomes: blood lipids (TC, TG, HDL-C, LDL-C, FFA, ApoA1, ApoB, ApoA1/ApoB, hs-CRP), 25 studies; glucose parameters (FBG, FBI, HbA1c, HOMA-IR, C-pep, 2hPPG, 2hPPI), 19 studies; anthropometric parameters (BW, BMI, WHR, BFP, TFA, VFA, SFA, WC, DBP, SBP) 20 studies; inflammatory cytokines (CRP, IL-6, IL-10, TNF-*α*), ten studies; and liver function tests, four studies. Impacts of BFF interventions on gut microbiota composition (relative abundance, correlation with genes/ clinical parameters) were investigated in four studies,[57,62,70,76] with only Akamine et al. [62] measuring plasma short-chain fatty acid levels as a functional outcome of microbiota changes.

### Study findings

Post-intervention, all 26 studies reported significant within-group changes in means of some target outcomes, compared to baseline. The p-values of significant (p<0.05) between-group differences for each study are shown in Online Supplemental material 4. Statistically significant impacts of BFFs on discrete cardiometabolic outcomes when compared to controls were observed in 19 studies.[53,56,58–61,63–69,71–74,76,77] Tenore et al. [76] and Akamine et al. [62] reported significant intergroup differences in gut microbiota composition. No significant difference between groups in any measured outcomes were found in five studies,[54,55,57,75,78] and one study [70] did not perform between-group analysis. No significant adverse effects were reported. To allow for suitable comparisons within BFF subtypes, the results are presented according to the major microorganisms involved in the production of each fermented food.

#### Lactofermented kimchi, red ginseng, ash kardeh and Annurca apple

Lactofermented foods are produced through lactic acid fermentation with LAB. These foods were used as interventions in eight studies: 10-day fermented kimchi vs. 1-day-fermented fresh kimchi;[53–57,73] fermented red ginseng vs. yeast placebo;[63,73,76] lactofermented Annurca apple puree;[76] and ash kardeh.[63] The target population was adults with T2DM in three studies;[54,63,73] overweight/ obesity in two studies;[53, 57] MetS in two studies;[55, 56] and cardiovascular risk factors in one study.[76] South Koreans were the target population in six studies,[53–57] with Tenore et al. [76] and Salehi et al. [63] conducted in Italian and Iranian populations respectively.

Blood lipid parameters were measured in seven studies.[53–57,63,73,76] Kim et al. [53] conducted a crossover trial of 300 g/d fermented (FeK) vs. fresh kimchi (FrK) in 22 participants with BMI ≥ 25kg/m². TC was significantly reduced in the FeK group compared to baseline; this change was significantly different from FrK (FeK -0.3 vs. FrK -0.1 mmol/L; p<0.05). However, in Han et al.’s [57] eight-week 180 g/d parallel study of a similar female population (n=24), HDL-C significantly improved in the FeK group compared to baseline, but this change was not significantly different from the FrK group. Notably, some subjects in both studies had elevated TG levels (≥1.7 mmol/L). In An et al. [54] and Lee et al. [56] (abstracts only, dosages unclear), eight-week crossover trials with FeK and FrK in participants with T2DM (n=20) or MetS (n=16) respectively, there were no reported significant between-group differences in blood lipids. Lactofermented ash kardeh (AK) was consumed for six weeks, along with routine diabetic treatment, by 48 participants with T2DM in an open label parallel trial;[63] many of the participants would fit MetS criteria (obese, abnormal TG and HDL-C levels, hypertension). In comparison to the routine diabetic treatment control group, 250 g/d AK consumption resulted in significant reductions in TC (AK -0.2 ± 0.5 vs. control 0.1 ± 0.3 mmol/L; p=0.025) and TG (AK -0.3 ± 0.3 vs. control -0.1 ± 0.2 mmol/L; p=0.003), as well as a significant increase in HDL-C (AK 0.04 ± 0.1 vs. control -0.05 ± 0.2 mmol/L; p=0.048). Consumption of 2.7 g/d fermented red ginseng (FRG) vs. placebo for four weeks by 42 T2DM/ prediabetic participants did not elicit any significant between-group differences in measured blood lipids,[73] although the FRG group had significant improvements in TC, HDL-C and LDL-C compared to baseline. Tenore et al. [76] compared consumption of 125 g unfermented Annurca apple puree (AAP) to lactofermented AAP (lfAAP) and a probiotic containing an identical dosage of the same LAB (3.0 × 10^8^ CFU/ g *Lacticaseibacillus rhamnosus* LRH11 and *L. plantarum* SGL07), in 90 Italian adults with cardiovascular risk factors. After 8 weeks, HDL-C was significantly increased in all groups compared to baseline; both lfAAP and AAP groups in relation to each other; and to the probiotic alone (lfAAP 0.6 vs. AAP 0.5 vs. LAB 0.2 mmol/L; p<0.05). Notably, there was a 31% drop-out rate. Overall, three included studies of lactofermented foods found significant (p<0.05) between-group differences in the blood lipid parameters TC, TG or HDL-C.

Glucose parameters were measured in all lactofermented food studies. Most participants in Kim et al.’s [53] study had elevated FBG at baseline (5.7 ± 0.6 mmol/L); fermented kimchi (FeK) significantly reduced FBG and FBI compared to baseline, but only the change in FBG was significantly different to fresh kimchi (FrK) (FeK -0.3 vs. FrK -0.2 mmol/L; p<0.05). In An et al.,[54] only the FrK group had improved HbA1c compared to baseline, whilst in An et al.,[55] both groups showed improvement in HbA1c, HOMA-IR and FBI. However, there were no significant between-group differences in either of these studies, or in Lee et al.,[56] Han et al. [57] or Tenore et al..[76] Ash kardeh (AK) [63] significantly reduced FBG levels compared to control (AK -1.1 ± 1.4 vs. control -0.2 ± 0.5 mmol/L; p=0.003). In Oh et al.,[73] the only significant changes from baseline were found in the FRG group (FBG, 2hPPG); after four weeks, the FRG group had significantly reduced 2h postprandial glucose (FRG -1.6 vs. placebo -0.5 mmol/L; p=0.008), and increased 2h postprandial insulin (FRG 17.1 vs. placebo -2.6 µU/mL; p=0.040) compared to placebo. Overall, three studies observed a significant reduction in the glucose parameters FBG or 2hPPG/ 2hPPI compared to control.

All lactofermented food studies collected anthropometric data, except for Oh et al..[73] Salehi et al. [63] found that ash kardeh (AK) significantly reduced SBP compared to the control group (AK - 13.47 ± 14.01 vs. control -0.43 ± 5.62 mmHg; p<0.001). In Kim et al.,[53] both groups had significant improvements in BW, BMI and BFP compared to baseline, whilst fermented kimchi (FeK) also reduced WHR. Compared to fresh kimchi (FrK), FeK consumption significantly reduced BFP (FeK -0.7 vs. FrK -0.3%; p<0.05), SBP (FeK -4.8 vs. FrK -3.7 mmHg; p<0.05), and DBP (FeK -4.2 vs. FrK -1.7 mmHg; p<0.05). Lee et al. [56] also showed that FeK significantly reduced DBP (FeK -3.7 mmHg vs. FrK not stated; p=0.037). Compared to baseline, An et al. [55] found that both the FeK and FrK groups had significant reductions in BW, BMI, WC and BFP, with only the FeK group having significant improvements in SBP and DBP. However, there were no significant intergroup differences. Similarly, Han et al. [57] had significant within-group improvements in WC, BFP, DBP (FrK) and SBP (FeK) but no significant between-group differences. An et al. [54] had significant within-group change in DBP in the FeK group but no significant between-group differences. Although all studies measured inflammatory markers, only Lee et al. [56] showed a significant between-group difference in CRP (FeK 208.5 µg/mL vs. FrK not stated; p=0.048). Overall, three studies reported significant between-group reductions in SBP and DBP compared to control, with one study reporting a significant reduction in CRP.

Han et al. [57] and Tenore et al. [76] collected faecal samples for assessment of changes in gut microbiota composition. In Han et al.,[57] faecal DNA pyrosequencing and blood RNA microarray were performed on ten and four randomly selected participants per group respectively. Identified bacterial taxa were also correlated with relevant obesity genes and clinical parameters. *Bifidobacterium* was significantly increased in both groups after eight weeks. In correlation analysis, significant negative correlations were found between *Actinobacteria* and BFP; *B. longum* and WC; *Bifidobacterium* and BW; and, *Bifidobacterium* and BMI. In the fermented kimchi group, the versican (regulates inflammation) and acyl-CoA synthetase long-chain family member 1 (regulates lipid metabolism) genes were significantly upregulated (p<0.05); these genes were found to negatively correlate with SBP and DBP (p<0.05). Tenore et al. [76] also examined changes in gut microbial composition, and found that when comparing lfAAP, AAP and a LAB probiotic, there were significant (p<0.05) increases in *Bifidobacterium* (AAP 7.7 × 10^5^ vs. lfAAP 5.0 × 10^4^ vs. probiotic 3.8 × 10^5^ CFU/mL) and LAB (AAP 1.1 × 10^6^ vs. lfAAP 3.5 × 10^3^ vs. probiotic 1.4 × 10^5^ CFU/mL); and, reductions in *Bacteroides* (AAP -4.9 × 10^3^ vs. lfAAP -1.8 × 10^3^ vs. probiotic -2.5 × 10^3^ CFU/mL) and *Enterococcus* (AAP -3.5 × 10^2^ vs. lfAAP -9.2 × 10^2^ vs. probiotic -3.4 × 10^3^ CFU/mL). The authors stated that “post follow up, lfAAP showed highest stability of efficacy, followed by LAB, & AAP”.

Overall, the results of the included lactofermented food studies were inconsistent. However, there was a trend of within-group improvements in measured metabolic parameters. Several significant between-group improvements were reported in two studies: Kim et al.’s [53] fermented kimchi study (SBP, DBP, BFP, FBG, TC), and Salehi et al.’s [63] ash kardeh study (SBP, FBG, TC, TG, LDL-C).

#### Chungkookjang, kochujang and doenjang

Soybean-based jangs fermented with *Bacillus* and *Aspergillus* species were used as interventions in six studies: chungkookjang,[65,67,77] kochujang [70, 72] and doenjang.[71] Shin et al. [65] investigated adults with impaired FBG, whilst the other five studies [67,70–72,77] were conducted in overweight/ obese adults (BMI ⩾23 kg/m² & WHR ⩾0.90 (M), ⩾0.85 (F)). All studies were conducted in South Korean adults.

All six studies measured blood lipids. Back et al. [67] showed that 12-week consumption of chungkookjang (CKJ) (26 g/d) significantly reduced ApoB compared to baseline, and in comparison to placebo (CKJ -0.2 vs. placebo -0.1 µmol/L; p=0.027; n=60). In Cha et al.,[72] the kochujang (KCJ) group (32 g/d) showed improvement in TG, ApoB, ApoB/ ApoA1 and ApoA1 after 12 weeks, whilst the placebo group had improvements in ApoA1 and ApoB. Compared to placebo, KCJ significantly reduced TG (KCJ -0.2 vs. placebo 0.1 mmol/L; p=0.049; n=60). In Byun et al.’s [77] (n=83; 35 g/d CKJ) 12-week crossover trial, there were changes from baseline in both the CKJ female (TG, ApoA1, ApoB) and male (TC, TG, ApoA1) groups, as well as in the placebo group. In Cha et al. [71] (n=51), both the doenjang (9.8 g) and placebo groups had improvements in TC, LDL-C, ApoB and ApoA1 after 12 weeks. However, there was no significant difference in any lipid measures between groups in either of these studies. These overweight/ obese study populations did not have any lipid abnormalities at baseline. Shin et al.,[65] an eight-week, three-arm parallel trial comparing 20 g/d of CKJ or red ginseng CKJ (RGCKJ) vs. a starch control in adults with impaired FBG, did show significant downward trends in TC and LDL-C (CKJ, RGCKJ), and ApoB/ApoA1, but did not find any significant between-group differences in blood lipids. However, in this study, 20 g of both CKJ and RGCKJ had significant within-group improvements in FBG, and when compared to control (CKJ -1.7 vs. RGCKJ -1.1 vs. control 0.2 mmol/L; p<0.05). Han et al. [70] conducted a six-week, three-arm parallel study in overweight/ obese adults comparing 19g/d high effective microorganisms traditional KCJ (n=19), low effective microorganisms traditional KCJ (n=18) and commercial KCJ (n=17). However, it appears that they did not conduct between-group analysis; they did find that significant within-group changes only occurred in the high effective microorganisms KCJ group (TC, LDL-C, HDL-C, TG). Cha et al. [72] found no significant differences in glucose parameters between KCJ and placebo groups; the remaining studies did not measure these parameters. Overall, although all studies found within-group differences in blood lipid parameters, two studies found significant between-group differences in TC or TG.

In Byun et al.,[77] chungkookjang (CKJ) significantly reduced WC (CKJ -1.05 cm vs. placebo 1.84 cm; p=0.0067), WHR (CKJ -0.01 vs. placebo 0.02; p=0.0083) and BFP (CKJ -0.84% vs. placebo 0.2%; p=0.0488) in female participants (n=43). After 12 weeks, Cha et al. [72] found that kochujang elicited a significant reduction in VFA (kochujang -4.8 cm^2^ vs. placebo -0.4 cm^2^; p=0.043). Cha et al. [71] showed that, compared to baseline, doenjang (DE) significantly reduced BW, BFM, BFP and VFA, whilst both DE and placebo groups had improvements in TFA, SFA and WHR. There was a significant between-group difference in BW (DE -0.8 kg vs. placebo -0.3 kg; p<0.001), BFM (DE-0.7 kg vs. placebo -0.4 kg; p<0.001), VFA (DE -8.6 cm2 vs. placebo -0.6 cm2; p=0.041) and BFP (DE -0.6% vs. placebo -0.4%; p=0.007). Han et al. [70] only reported significant within-group differences: high effective microorganisms kochujang reduced WC and VFA; commercial kochujang reduced WC and WHR. This study also measured “beneficial, harmful and other” faecal microorganisms, but found no significant within-group differences. Back et al.’s [67] 12-week administration of 26 g/d CKJ did not elicit any significant within- or between-group differences in anthropometric measures. Overall, two studies found significant reductions in certain anthropometric parameters compared to control, with another only finding significant differences in the women’s subgroup.

Despite significant differences from baseline in lipid and glucose parameters, jangs elicited few consistent between-group improvements. However, several studies showed small but significant changes in anthropometric parameters in overweight/ obese participants: BW, BFM, BFP, WC, WHR and VFA.

#### Shiokoji, miso, touchi, amazake and kochujang

*Aspergillus oryzae*-fermented products were administered to different target populations in five studies: Touchi-extract supplemented Houji tea [66] and shiokoji [78] in adults with mild hyperglycaemia or borderline/ mild T2DM; *A. oryzae*-fermented KCJ in adults with mild hyperlipidaemia;[74] brown rice amazake vs. white rice amazake in adults with MetS;[62] and miso in adults with high normal BP or stage I hypertension.[68] South Korean adults were participants in three studies, with Japanese adults investigated in Akamine et al. [62] and Lim et al..[74]

Nakamura et al. [78] administered 4 weeks of 15 g/d of shiokoji or placebo to 49 adults with mild hyperglycaemia (FBG 5.6–6.9 mmol/L). After data was collected, two analyses were conducted: primary analysis (n=47), and secondary analysis (n=30), for which participants who had finished the study were deemed ineligible due to marked changes in lifestyle (alcohol consumption/ exercise) and/ or HOMA > 5.0 post food allocation. In the primary analysis, there was no significant difference between the groups. In the secondary analysis, shiokoji significantly reduced FBG (shiokoji -0.2 mmol/L vs. placebo 0.1 mmol/L; p=0.02) after 4 weeks. However, after 12 weeks, there was no significant difference between the groups. Blood lipids and anthropometric parameters were collected but not reported. To 38 borderline/ mild T2DM subjects, Fujita et al. [66] administered 0.3g *Aspergillus*-fermented Touchi extract supplemented with Houji tea (TE) vs. control for 12 weeks. The TE group had within-group improvements in FBG and HbA1c. Compared to the control, TE significantly improved FBG (TE -0.5 mmol/L vs. control 0.2 mmol/L; p<0.05) and HbA1c (TE - 0.5% vs. control 0.1%; p<0.01). There were no significant between-group differences in anthropometric parameters or blood lipids.

In Lim et al.’s [74] 12-week parallel study, 30 adults with mild hyperlipidaemia (LDL-C 2.8–4.9 mmol/L or TC 5.2–6.7 mmol/L) were administered 34.5 g/d of *A. oryzae*-fermented kochujang (KCJ). The KCJ group had a significant reduction in TC compared to baseline, as well as compared to placebo (KCJ -0.5 vs. placebo -0.2 mmol/L; p=0.045). No significant between-group differences in anthropometric parameters were reported.

Kondo et al. [68] studied 40 adults with high-normal BP (130-139/85-89 mmHg) or untreated stage I hypertension (140-159/90-99 mm Hg), who were given a 2:1 mixture of high-ACE inhibitory MK-34-1 miso and Nenrin (common) miso, or a soy food control for 8 weeks. The miso group had significant improvements in LDL-C, night-time SBP and DBP compared to baseline, but there were no other within- or between-group differences in glucose, lipid or liver function parameters. There were significant between-group changes in BW (miso 0.636 kg vs. control 0.073 kg; p<0.05). In the whole group, as well as the stage I hypertensive subgroup, miso was found to significantly (p<0.05) decrease night-time SBP and DBP when compared to the control; it did not significantly affect daytime blood pressure.

Akamine et al. [62] compared the 4-week consumption of 350 g/d brown rice amazake (BA) to a white rice amazake (WA) control in 40 adults with MetS (BMI ⩾25 kg/m² and any 2 of: (1) TG ⩾1.7 mmol/L; (2) HDL-C ⩽1.0 mmol/L; (3) SBP ⩾130 mmHg +/-DBP ⩾85 mmHg; or (4) FBG ⩾5.6 mmol/L). There were no significant within- or between-group differences in any glucose, lipid or anthropometric parameters, or short-chain fatty acid plasma concentration. The primary outcome of this study was changes in gut microbiota composition. The WA group had a significant increase in *Flavonifractor plautii* but there were no other within-group differences. Comparing the two groups, after four weeks intervention, there were significant increases in the BA group in: the family *Porphyromonadaceae* (p=0.013); the genera *Parabacteroides* (p=0.011), *Butyricicoccus* (p=0.012) and *Sutterella* (p=0.001); and species *Sutterella wadsworthensis* (p=0.001), *Lactobacillales bacterium DJF B280* (p=0.005), *Firmicutes* bacterium DJF VP44 (p=0.038) and *Eubacterium* spp. A2 207 [84] (p=0.012). There were no significant differences between the groups in alpha diversity, beta diversity or taxonomy-based abundance analyses at the phylum level. To assess whether these gut microbial changes correlated with metabolic biomarkers in each subject at species level, correlation analyses were carried out. Significant correlations (correction r>0.4 or r<-0.4; p<0.05) were identified among the included 41 taxa and 13 clinical indices. Significant negative correlations were found between *Sutterella wadsworthensis*/ blood glucose (p=0.032); *Lactobacillales* bacterium DJF B280/ TG (p=0.006); and *Eubacterium* spp. A2 207/ TG (p=0.001). Significant positive correlations were found between *Lactobacillales bacterium* DJF B280/ HDL-C (p=0.037), and *Eubacterium* spp. A2 207/ LDL-C (p=0.046).

Overall, *A. oryzae*-fermented foods elicited few consistent, significant within- or between-group

#### Tempeh

Tempeh (*Rhizopus* spp.) products were used as interventions in three studies: Wirawanti et al.’s [58] tempeh drink study in hypercholesterolaemic adults; Afifah et al.’s [59] study in women with hyperlipidaemia; and Nadia et al.’s [60] study of women with premenopausal obesity. All tempeh studies were conducted with Indonesian participants.

In Nadia et al.’s [60] open, parallel study of tempeh gembus in an isocaloric diet (30 kcal/kg of BW/d) vs. isocaloric diet alone, all 40 participants were prediabetic or diabetic (range 2.6 to 5.1 mmol/L; median ∼ 3.2 mmol/L), with BMI between 25.1 to 47.7kg/m². At baseline, most participants in this study had low HDL-C (⩽1.0 mmol/L) and elevated TG (⩾1.7 mmol/L) levels. After four weeks, 150 g/d of tempeh gembus significantly improved FBG, FBI, HOMA-IR, TG, HDL-C and hs-CRP in both groups. Between-group differences were observed in FBG (tempeh -2.3 vs. control -0.4 mmol/L; p=0.00); HOMA-IR (tempeh -18.4 vs. control -0.6; p=0.00) and FBI (tempeh -14.2 µIU/L vs. control -0.7 µIU/L; p=0.00). TG (tempeh -0.1 vs. control 0.03 mmol/L; p=0.01) and hs-CRP (tempeh -1.94 mg/L vs. control -1.65 mg/L; p=0.03) and HDL-C were significantly improved by tempeh gembus consumption (tempeh 0.2 vs. control 0.2 mmol/L; p=0.00) compared to diet alone.

Afifah et al. [59] ran a 2-week open, parallel study in 45 women with hyperlipidaemia (TC ⩾5.2 mmol/L &/or LDL-C ⩾2.6 mmol/L). Tempeh gembus at two different dosages (103 g T1 and 206 g T2) and the control all showed improvements in TC and LDL-C compared to baseline. T1 and T2 significantly decreased LDL-C (T1 -1.1 vs. T2 -1.3 vs. control -0.7 mmol/L; p=0.035) when compared to each other and to the control (10 mg simvastatin); post-hoc analysis showed that T2 had the largest effect, significantly reducing LDL-C (means difference -0.6 mmol/L; p=0.011) when compared to control.

Hypercholesterolaemic (TC ⩾5.2 mmol/L; n=54) adults were provided tempeh drinks made with local (TA) or imported (TB) soybeans, or no intervention, in Wirawanti et al.’s [58] open, parallel study. Both TA and TB were found to significantly decrease TC after four weeks, and the no-intervention control group had reductions in LDL-C and HDL-C. TA and TB improved LDL-C (TA -0.1 vs. TB -0.03 vs. control 0.2 mmol/L; p=0.01) compared to control; there was no significant difference between TA and TB.

Compared to controls, even over very short periods of time (i.e. two to four weeks), tempeh seemed to significantly improve blood lipids (LDL-C, HDL-C, TG) in hyperlipidaemic participants, and improved FBG, HOMA-IR and FBI in a prediabetic/ diabetic obese female population.

#### Other BFFs

A variety of other BFFs were administered to different populations in four studies: Kim et al. (2011a) administered brown rice lees (LB, with *A. oryzae, R. oryzae,* LAB, yeasts) to adults with T2DM; Somanah et al. [61] examined the effect of fermented papaya preparation (FPP, with *E. faecalis, A. oryzae*) in adults with neodiabetes; Jung et al. [69] used a yeast-fermented soy powder (Q-Can) in adults with cardiovascular risk factors; and, Mohammadi et al. [75] used fermented garlic extract within chocolate in adults with hypertension.

Somanah et al. ’s [61] 14-week parallel fermented papaya preparation (FPP) (6 g) vs. water control study (n=127) yielded some within-group improvements (control males and females: TC, LDL-C, HDL-C; control males: SBP, DBP; FPP (M): TG; FPP females: BMI, DBP), but no significant between-group differences in glucose, lipid or anthropometric parameters in Mauritian adults with neo diabetes (FBG 5.1–5.9 mmol/L). The only significant finding was a reduction in CRP (p=0.018, inconsistent data provided) in the FPP group compared to the control. It should be noted here that this study was assessed as being of poor quality with a high risk of bias, due to a high number of dropouts (27), substantial differences in group size allocation and unexplained, inconsistent numbers of participants with missing outcome data for each outcome.

Kim et al. [64] administered 40 g brown rice lees (LB), a by-product of Korean rice wine, for 12 weeks to 30 South Korean adults with T2DM (FBG ⩾7 mmol/L or ⩾11.1 mmol/L in OGTT), finding no significant change in any glucose parameters compared to the 45 g mixed grain (MG) control. The MG group had significant within-group improvements in TC, LDL-C, SBP, AST and ALT, whilst the LB group had improved WC. Compared to MG, LB significantly increased TC (LB 0.3 vs. MG -0.2 mmol/L; p<0.05) and LDL-C (LB 0.2 vs. MG -0.3 mmol/L; p<0.05). The authors explained the unexpected increase in these parameters as likely due to “relatively high-fat content” of LB and “high insoluble fibre content” of MG, resulting in “an increase in cholesterol levels in the LB group vs the MG group”. In the LB group, there were also significant between-group improvements in TC (LB -2.8 cm vs. MG -0.9 cm; p<0.05), as were AST (LB -4.5 IU/mL vs. MG -0.1 IU/mL; p<0.05) and ALT (LB -6.7 IU/mL vs. MG 0.1 IU/mL; p<0.05).

In a 12-week crossover trial, Jung et al. [69] compared a yeast-fermented soy powder (FS), Q-Can, to a germinated brown rice (GBR) powder control in 29 American adults with cardiovascular risk factors (⩾ 2: smoking; SBP ⩾140/90 mmHg; LDL-C ⩾2.9 mmol/L; HDL-C ⩾1.0 mmol/L; TG ⩾1.7 mmol/L; FBG ⩾6.1 mmol/L; BMI ⩾25 kg/m²; family history of heart disease). Compared to baseline, FS significantly improved TC, LDL-C and ApoB, whilst GBR had improvements in HDL-C. Between groups, there were significant improvements in TC (FS -0.23 mmol/L vs. GBR 0.14 mmol/L; p=0.0024), LDL-C (FS -0.18 mmol/L vs. GBR 0.4 mmol/L; p=0.0317), HDL-C (FS -0.03 mmol/L vs. GBR 0.09 mmol/L; p=0.0036) and ApoA1 (FS -1.4 µmol/L vs. GBR 1.4 µmol/L; p=0.0390). There were no significant between-group differences in anthropometric measures or measured glucose parameters.

Mohammadi et al. [75] administered dark chocolate containing 650 mg of fermented garlic extract or a dark chocolate control to 44 hypertensive (SBP ⩾130 mmHg ± DBP ⩾80 mmHg, or consuming antihypertensive drug) adults in a 6-week crossover trial. No significant differences were found within- or between-groups in lipid, anthropometric or inflammatory parameters. Glucose metabolism was not investigated.

## DISCUSSION

Our review found that almost all of the studied BFFs, compared to controls, significantly improved certain lipid, glucose, anthropometric and inflammatory parameters in those with overweight/ obesity, MetS or T2DM. Consumption of some BFFs led to significant beneficial changes in gut microbiota composition, with correlations to metabolic outcomes and gene expression. However, the results were inconsistent across studies, even within BFF subgroups such as fermented kimchi, and 46% of the studies were at medium to high risk of bias. The clinical relevance of these results is yet to be determined and should consider the methodological limitations of these studies.

The strength of our review lies in our robust and inclusive methodology. We were able to locate a culturally diverse range of relevant RCTs from 2001 onwards by ensuring our review had no language limits, searched from inception and included specific traditional BFF terminology from many cultures. To recognise and assess the emerging role of the gut microbiota in nutritional research, we included changes in gut microbiota composition as one of our target outcomes. To the best of our knowledge, our included studies have not been assessed in other systematic reviews or meta-analyses, and this is the first review using systematic Cochrane methodology to identify the impact of BFFs (excluding red yeast rice and vinegar) in our target population. However, our ability to perform meta-analyses, and the extrapolation of our findings, are limited by the diversity of BFFs, undeclared microbial strains and counts, dosage, intervention length and administration, and variable target conditions and study populations.

To the best of our knowledge, our systematic review is unusual in collecting data on changes in gut microbiota composition with cardiometabolic outcomes. Colonic microbiota has an established role in the development of obesity, MetS and T2DM, and fermented food consumption affects gut microbiota composition. However, only 15% (4/26) of our reviewed studies collected faecal samples for gut microbiota profiling,[57,62,70,76] with only Han et al. [57] and Akamine et al. [62] correlating these findings to gene expression and/or metabolic outcomes. Kochujang with varying levels of effective microorganisms did not elicit significant changes in gut microbiota composition.[70] In the other studies, BFF consumption generally increased the abundance of beneficial bacteria. Both fresh and fermented kimchi significantly increased *Bifidobacterium*,[57] but there were no between-group differences. Lactofermented Annurca apple puree, compared to a probiotic or nonfermented puree control, significantly increased beneficial *Bifidobacterium* and LAB, whilst reducing potential pathogens *Bacteroides* and *Enterococcus*.[76] A similar trend was observed in Akamine et al. [62] where brown rice amazake significantly increased beneficial bacteria on a family and genus level, including *Sutterella wadsworthensis*, *Lactobacillales* bacterium DJF B280, *Firmicutes* bacterium DJF VP44 and *Eubacterium* spp. A2 207, compared to white rice amazake. These findings concur with BFF-induced increases in beneficial bacteria and concurrent reductions of pathogenic bacteria during *in vitro* and *in vivo* studies,[85–87] as well as a limited number of human interventional trials.[88–91] Additionally, both Han et al. [57] and Akamine et al. [62] correlated taxa with clinical indices, finding significant negative and positive correlations indicating the potential beneficial effects of certain bacteria on discrete metabolic outcomes. However, the included studies suffered from short intervention periods and used a variety of methods to quantify microbes. Interpretation of these results must consider the effects of short- and long-term dietary changes on gut microbiota,[92] and the relative paucity of gut microbial data in such studies. Due to their important role in diet and metabolism, we suggest that all RCTs of fermented foods should collect data on the gut microbiota,[93] including microbial metabolite levels, and correlations with metabolic outcomes and gene expression.

In comparing our review with similar studies, we found Gille et al.’s [18] 2018 review of meta-analyses of fermented foods and noncommunicable disease, and SaeidiFard et al.’s [94] 2020 meta-analysis of fermented foods and inflammation. Notably, the studies included in these reviews did not overlap with our included publications. Gille et al.’s review stated that “The literature on fermented plants is characterized by a wealth of *in vitro* data, whose positive results are not corroborated in humans due to the absence of RCTs”.[18] Their section on “fermented foods of plant origin” focused only on those relevant to the Swiss population (coffee, wine, beer, sauerkraut, fermented olives). SaeidiFard et al.’s [94] meta-analyses included healthy participants, fermented dairy products and probiotic-added nonfermented foods. They found that CRP and IL-6 were not improved by fermented food intake, with only a reduction in TNF-α. Although nine of our included studies [53–57,61,70,75] measured some inflammatory markers (including CRP, IL-10, IL-6, TNF-a), only one fermented vs. fresh kimchi study [56] (p=0.048) and the partly biased fermented papaya study [61] (p=0.018) observed significant reductions in CRP compared to controls. We also identified several recent meta-analyses of RCTs using vinegar or red yeast rice/ monacolin/ xuezhikang in adults with obesity, T2DM or dyslipidaemia. All the meta-analyses of vinegar consumption found that glycaemic control was improved in individuals with IGT or T2DM, including: FBG;[42, 44] HbA1c;[40, 42] and acute glucose response/ postprandial glucose and insulin.[41, 45] Cheng et al.[42] also found statistically significant improvement in LDL-C and TC levels. However, there was substantial heterogeneity reported across studies within each meta-analysis, and there were varying numbers of RCTs assessed by each review. Like vinegar, many BFFs contain acetic acid and short chain fatty acids. Out of 19 studies which measured glucose outcomes, our review found that only six studies [53,60,64–66,73,75] of different BFFs had significant improvements in some aspect of glycaemic control (FBG, HbA1c, FBI, 2hPPG and 2hPPI). Importantly, BFF subgroups, such as fermented kimchi, did not have the same effects on glycaemic control across all studies, which may be due to differences in study length, dosage and study populations. Meta-analyses of red yeast rice identified significant improvements in LDL-C, TC and TG compared to placebo in dyslipidaemic patients.[36,37,95] Our review also found that some lactofermented foods,[53,63,76] jangs,[67,72,74] tempeh,[58–60] brown rice lees [64] and fermented soy powder [69] significantly improved lipid parameters compared to controls. Furthermore, while some anthropometric parameters (BFP, BFM, BW, WC, WHR, VFA) were significantly improved in six studies,[53,64,68,71,72,77] results were inconsistent across and within BFF subgroups. As we did not conduct meta-analyses, these comparisons with other reviews need to be made with caution.

For several reasons, caution must be exercised by clinicians and policymakers when considering our review’s findings. The findings from nutritional clinical trials may not be amenable to meta-analysis by their very nature; unlike drug trials, methodology varies considerably across studies, and dietary intake is notoriously difficult to assess and control.[96] For example, consider the fresh vs. fermented kimchi subgroup of studies, performed by the same clinical group.[53–57,73] Kim et al. [53] supplied a larger dose for a shorter period (300 g/d for 4 weeks) than Han et al. [57] (180 g/d for 8 weeks) in a similar obese population. However, only Kim et al. [53] found significant between-group differences in FBG, TC, BFP, SBP and DBP post intervention. An et al. [55] also studied a prediabetic population with MetS, administering 300 g/d over eight weeks but, like Han et al.,[57] did not find any significant between-group differences. In relation to the role of diet in nutritional RCTs, several of the studies we assessed implemented isocaloric diets or provided meals in addition to the BFF/ placebo for the duration of the study. Although these were controlled for, these dietary changes may have confounding effects on study outcomes.[97] Small sample sizes may affect statistical power in some of the included studies. Three studies only recruited women [57,59,60] and two found significant differences only in the male or female subgroups:[61, 77] clinically speaking, cardiovascular risk may vary between genders in those with MetS.[98] Ten included studies [53–62] were at risk of selection bias as they did not provide sufficient information regarding random sequence generation and allocation concealment. Notably, 21 out of 26 included studies were conducted in East Asian populations (South Korea, Japan, Indonesia) with one from Western Asia [63] (Iran) using traditional BFFs from these regions, which reflects the importance of these products in their diets. The other studies from Italy,[76] the USA,[69] Iran [75] and Mauritius [61] used non-traditional fermented interventions. East Asians may be at higher risk of poor cardiometabolic outcomes at a lower BMI than Caucasians.[99] This is reflected in our decision to include overweight/ obese participant studies with a BMI cut off of ⩾23 kg/m², and MetS waist circumference cut off for East Asians as per the WHO [100] obesity guidelines (Online Supplemental material 1). Our review indicates the need for consensus on consistent and robust study designs for BFFs, including dosage studies, consistent comparators, longer intervention periods and larger participant pools.

Our systematic review indicates that BFFs have the potential to improve metabolic outcomes in individuals with obesity, MetS and T2DM. The marginal effects of BFFs may be linked to the lack of qualified systematic studies. Based on existing results, we suggest that BFFs may be best used in conjunction with lifestyle modifications and appropriate medications. However, the paucity of studies in this area, in contrast to the diverse range of available BFFs, suggests that large-scale, long-term and well-designed RCTs with a low risk of bias are required before more definitive conclusions can be made.

## Data Availability

All data produced in the present work are contained in the manuscript

## AUTHOR CONTRIBUTIONS

MC, EE and KH conceived the study. MC, EE, KH and LJ contributed to planning of the review and determination of its scope. MC and KH designed the protocol with input from HB, EE, NL and LJ. MC and HB developed the search strategy and executed the search, with HB translating the searches for multiple databases. MC and NL screened the studies, extracted data, assessed quality of evidence and performed data analysis. MC drafted and revised the manuscript, with editorial input from KH, EIE, LJ, NL and HB. All authors read and approved the final manuscript and order of authorship.

