## Supplemental material 1 for "The impact of botanical fermented foods on obesity, metabolic syndrome and type 2 diabetes: a systematic review of randomised controlled trials"

**Metabolic syndrome diagnostic components**

| Elevated waist circumference* or elevated waist-to-hip ratio** | WC: European/ North American M: >102 cm; F: >88 cm Asian/ Central & South America M: >90 cm; F: >80cm Middle East/ Med./ Africa M: >94 cm; F: >80 cm  WHR: M: >0.9; F: >0.85 |
| --- | --- |
| Elevated triglycerides *or treatment for this lipid abnormality* | ≥150 mg/dL (1.7 mmol/L) |
| Reduced HDL-cholesterol *or treatment for this lipid abnormality* | M: <40 mg/dL (1.0 mmol/L) F: <50 mg/dL (1.3 mmol/L) |
| Elevated blood pressure *or treatment for hypertension* | Systolic ≥130 and/or diastolic ≥85 mm Hg |
| Elevated fasting glucose or diagnosed T2DM *or treatment for elevated glucose* | ≥100 mg/dL (5.5 mmol/L) |

Adapted from Alberti et al.,[1,2] *WHO,[3] and **WHO.[4] T2DM, type 2 diabetes mellitus; Med., Mediterranean.

1	Alberti K, Eckel RH, Grundy SM, *et al.* Harmonizing the metabolic syndrome. *Circulation* 2009;**120**:1640–5. doi:10.1161/CIRCULATIONAHA.109.192644

2	Alberti KGMM, Zimmet P, Shaw J. Metabolic syndrome—a new world-wide definition. A Consensus Statement from the International Diabetes Federation. *Diabet Med* 2006;**23**:469–80. doi:10.1111/j.1464-5491.2006.01858.x

3	WHO Consultation on Obesity (1999: Geneva S, Organization WH. Obesity : preventing and managing the global epidemic : report of a WHO consultation. World Health Organization 2000. https://apps.who.int/iris/handle/10665/42330 (accessed 29 Oct 2022).

4	World Health Organization. Definition, diagnosis and classification of diabetes mellitus and its complications : report of a WHO consultation. Part 1, Diagnosis and classification of diabetes mellitus. World Health Organization 1999. https://apps.who.int/iris/handle/10665/66040 (accessed 29 Oct 2022).
