## Supplemental material 2 for "The impact of botanical fermented foods on obesity, metabolic syndrome and type 2 diabetes: a systematic review of randomised controlled trials"

**Search strategy for MEDLINE**

1. randomised controlled trial.pt.

2. controlled clinical trial.pt.

3. (randomized or randomised).ab.

4. placebo.ab.

5. drug therapy.fs.

6. randomly.ab.

7. trial.ab.

8. groups.ab.

9. 1 or 2 or 3 or 4 or 5 or 6 or 7 or 8

10. exp animals/not humans.sh.

11. 9 not 10

12. metabolic syndrome/

13. diabetes mellitus, type 2/or diabetes mellitus, lipoatrophic/

14. hypertension/

15. insulin resistance/

16. INSULIN/

17. blood glucose/

18. blood pressure/

19. cholesterol, HDL/

20. cholesterol, LDL/

21. non-alcoholic fatty liver disease/

22. dyslipidemias/

23. PREDIABETIC STATE/

24. obesity/ or obesity, abdominal/ or obesity, morbid/

25. overweight/

26. 12 or 13 or 14 or 15 or 16 or 17 or 18 or 19 or 20 or 21 or 22 or 23 or 24 or 25

27. (metabolic syndrome* or metabolic disorder* or MetS or dyslipidemia* or dysglycemia* or hypertension or diabetes or prediabetes or neo diabetic or obesity or overweight or insulin or hyperlipidemia* or lipid or blood pressure or NAFLD or non-alcoholic fatty liver or microbiota or microbiome or microflora or flora or intestinal or dysbiosis or inflamm*).mp.

28. 26 or 27

29. FERMENTATION/

30. fermented foods/

31. (monascus or monacolin or red yeast rice or Korean diet).mp.

32. (fermented or fermentation).mp.

33. 29 or 30 or 31 or 32

34. 28 and 33

35. 11 and 34

**Search strategy for EMBASE**

1. metabolic syndrome X/

2. non insulin dependent diabetes/ or lipoatrophic diabetes/

3. hypertension/

4. insulin resistance/

5. insulin/

6. glucose blood level/

7. blood pressure/

8. high density lipoprotein cholesterol/

9. low density lipoprotein cholesterol/

10. nonalcoholic fatty liver/

11. dyslipidemia/

12. impaired glucose tolerance/

13. obesity/ or abdominal obesity/ or morbid obesity/ or diabetic obesity/

14. 1 or 2 or 3 or 4 or 5 or 6 or 7 or 8 or 9 or 10 or 11 or 12 or 13

15. metabolic syndrome* or metabolic disorder* or MetS or dyslipidemia* or dysglycemia* or hypertension or diabetes or prediabetes or neo diabetic or obesity or overweight or insulin or hyperlipidemia* or lipid or blood pressure or NAFLD or non-alcoholic fatty liver or microbiota or microbiome or microflora or flora or intestinal or dysbiosis or inflamm*).mp.

16. 14 or 15

17. fermentation/

18. fermented product/

19. (monascus or monacolin or red yeast rice or Korean diet).mp.

20. (fermented or fermentation).mp.

21. 17 or 18 or 19 or 20

22. 16 and 21

23. randomized controlled trial/

24. controlled clinical study/ or Random$.ti,ab. or randomization/ or intermethod comparison/ or placebo.ti,ab. or (compare or compared or comparison).ti. or ((evaluated or evaluate or evaluating or assessed or assess) and (compare or compared or comparing or comparison)).ab. or (open adj label).ti,ab. or ((double or single or doubly or singly) adj (blind or blinded or blindly)).ti,ab. or double blind procedure/ or parallel group$1.ti,ab. or (crossover or cross over).ti,ab. or ((assign$ or match or matched or allocation) adj5 (alternate or group$1 or intervention$1 or patient$1 or subject$1 or participant$1)).ti,ab. or (assigned or allocated).ti,ab. or (controlled adj7 (study or design or trial)).ti,ab. or (volunteer or volunteers).ti,ab. or human experiment/ or trial.ti.

25. 24 not 23

26. (((((((((((random$ adj sampl$ adj7 ("cross section$" or questionnaire$1 or survey$ or database$1)).ti,ab. not (comparative study/ or controlled study/ or randomi?ed controlled.ti,ab. or randomly assigned.ti,ab.)) or Cross-sectional study/) not (randomized controlled trial/ or controlled clinical study/ or controlled study/ or randomi?ed controlled.ti,ab. or control group$1.ti,ab.)) or (((case adj control$) and random$) not randomi?ed controlled).ti,ab. or (Systematic review not (trial or study)).ti. or (nonrandom$ not random$).ti,ab. or "Random field$".ti,ab. or (random cluster adj3 sampl$).ti,ab. or (review.ab. and review.pt.)) not trial.ti.) or "we searched".ab.) and (review.ti. or review.pt.)) or "update review".ab. or (databases adj4 searched).ab. or (rat or rats or mouse or mice or swine or porcine or murine or sheep or lambs or pigs or piglets or rabbit or rabbits or cat or cats or dog or dogs or cattle or bovine or monkey or monkeys or trout or marmoset$1).ti.) and animal experiment/) or Animal experiment/) not (human experiment/ or human/)

27. 25 not 26

28. exp experimental organism/ or animal tissue/ or animal cell/ or exp animal disease/ or exp carnivore disease/ or exp bird/ or exp experimental animal welfare/ or exp animal husbandry/ or animal behavior/ or exp animal cell culture/ or exp mammalian disease/ or exp mammal/ or exp marine species/ or nonhuman/ or animal.hw.

29. 28 not human/

30. 23 not 29

31. 27 or 30

32. 22 and 31

33. remove duplicates from 32

**Search strategy for Google Scholar**

"metabolic syndrome" OR MetS OR diabetes OR diabetic OR hypertension OR "insulin resistance" OR "blood pressure" OR cholesterol OR "non-alcoholic fatty liver" OR prediabetes OR obesity OR overweight OR microflora OR microbiota OR microbiome fermentation OR fermented OR ferment AND -milk

**Search strategy for Cochrane CENTRAL**

1. MesH descriptor: [Metabolic Syndrome] this term only

2. MesH descriptor: [Diabetes Mellitus, Type 2] this term only

3. MesH descriptor: [Diabetes Mellitus, Lipoatrophic] this term only

4. MesH descriptor: [Hypertension] this term only

5. MesH descriptor: [Insulin Resistance] this term only

6. MesH descriptor: [Insulin] this term only

7. MesH descriptor: [Blood Glucose] this term only

8. MesH descriptor: [Blood Pressure] this term only

9. MesH descriptor: [Cholesterol, HDL] this term only

10. MesH descriptor: [Cholesterol, LDL] this term only

11. MesH descriptor: [Non-alcoholic Fatty Liver Disease] this term only

12. MesH descriptor: [Dyslipidemias] this term only

13. MesH descriptor: [Prediabetic State] this term only

14. MesH descriptor: [Obesity, Abdominal] this term only

15. MesH descriptor: [Obesity] this term only

16. MesH descriptor: [Obesity, Morbid] this term only

17. MesH descriptor: [Overweight] this term only

18. {OR #1-#17}

19. (metabolic syndrome* OR metabolic disorder* OR MetS OR dyslipidemia* OR dysglycemia* OR hypertension OR diabetes OR prediabetes OR neo diabetic OR obesity OR overweight OR insulin OR hyperlipidemia* OR lipid OR blood pressure OR NAFLD OR non-alcoholic fatty liver OR microbiota OR microbiome OR microflora OR flora OR intestinal OR dysbiosis OR inflamm*):ti,ab,kw

20. #18 OR #19

21. MeSH descriptor: [Fermentation] this term only

22. MeSH descriptor: [Fermented Foods] this term only

23. (monascus OR monacolin OR red yeast rice OR Korean diet OR fermented OR fermentation):ti,ab,kw

24. {OR #21-#23}

25. #20 AND #24
