## Supplemental material 3 for "The impact of botanical fermented foods on obesity, metabolic syndrome and type 2 diabetes: a systematic review of randomised controlled trials"

**Online supplemental material 3**

**Production of botanical fermented foods**

Kimchi, a traditional Korean fermented food, comes in many regional variations. Generally, it is made by fermenting a mixture of Napa cabbage with salt, typically with added hot red pepper powder, green leeks, garlic, ginger, starch syrup, fermented seafood sauces and sometimes fruit or other vegetables.[1] Lactic acid bacteria (LAB) such as *Lactiplantibacillus plantarum* (formerly *Lactobacillus plantarum*) dominate the fermentation process. In our included kimchi studies,[1–5] fresh kimchi was stored for one day at 1°C, whilst fermented kimchi was stored for 5 hours at room temperature, then 10 days at 5–10°C.[1]

Fermented red ginseng was produced by 15-day fermentation of alcohol-extracted red ginseng with *L. plantarum* at 35-40°C.[6]

Lactofermented Annurca apple puree was produced by fermenting Annurca (*M. pumila* Miller cv Annurca) apple puree with *Lacticaseibacillus rhamnosus* (formerly *Lactobacillus rhamnosus*) LRH11 and *L. plantarum* SGL07 for 48 hours at room temperature.[7]

Ash kardeh is a traditional Iranian fermented food containing primarily LAB. It is prepared by grinding the Kardeh plant (*Biarum bovei Blume*) with rice, adding a small amount of flour-buttermilk/ yogurt starter and water. This mixture is fermented for 24 hours at room temperature, then boiled.[8]

Jangs are traditional Korean fermented soybean pastes. Chungkookjang is produced by fermenting unsalted boiled soybeans with *Bacillus subtilis* or *Bacillus licheniformis* from rice straw for two to three days.[9] Kochujang is made by mixing boiled soybeans with meju (boiled soybean bricks fermented for 20 to 60 days outdoors, attracting *Aspergillus* and *Bacillus* species), salt, malt-digested rice syrup, rice flour and red peppers.[10] Doenjang is also made with meju, but only contains a salt brine.[11] Both doenjang and kochujang are fermented for six months or more.

One included study [12] that compared chungkookjang to red ginseng chungkookjang used a non-traditional method of production. Cooled, steamed soybeans were inoculated with *B. subtilis* and fermented for 20 hours at 40°C. For red ginseng chungkookjang, concentrated red ginseng extract was added before fermentation.

*Aspergillus oryzae-*fermented kochujang was prepared as follows: koji was prepared by soaking, draining, steaming and cooling non-glutinous rice powder, before incubating it with *A. oryzae* for 48 hours at 30°C. This koji was then mixed with soybeans, red pepper powder, starch syrup, oligosaccharides, salt and water before fermenting for 30 days at 30°C.[13]

Koji, a mould commonly used in Japan, is produced by fermenting rice that has been soaked, drained and steamed with *A. oryzae* for up to 72 hours at room temperature. To make shiokoji or amazake, koji is mixed with salt or water respectively, before further fermentation of up to 14 days at room temperature.[14] In one included study,[15] white and brown rice amazake were made by soaking and steaming rice before adding *A. oryzae* and fermenting for 41 to 46 hours at 33 to 35C. Water was then added, with the mixture heated to 75 to 85C. After cooling to 50 to 70C, rice koji was mixed in, and maintained at this temperature for 6 to 12 hours. To make miso, koji and salt are added to a variety of cooked ingredients, most commonly soybeans and rice, then fermented for varying amounts of time. The 12% salt miso used in our included study[16] consisted of a 2:1 mix of Nenrin miso (70% malted rice, contains yeast, fermented for 46 days), and MK-34-1 miso (50% malted rice, no yeast, fermented for 5 days).[17]

Touchi is a traditional Chinese salt-fermented black soybean condiment. Production requires two steps: fungal fermentation with koji (*A. oryzae*), then salt brine fermentation by bacteria and yeast for more than six months.[18]

Tempeh, a traditional Indonesian staple, is usually produced by fermenting cooked soybeans with the filamentous fungi *Rhizopus oligosporus* or *Rhizopus oryzae* for 24 to 36 hours at 30°C. In one of our included studies,[19] germinated soybeans were used to make tempeh; germination involved soaking soybeans in water for 6 hours, leaving them covered at room temperature for 28 hours, with water application every 3 hours. Tempeh gembus was used as an intervention in two included studies,[20,21] and is made using soy pulp, a waste product of tofu production, as a substrate for 36-hour *R. oligosporus* fermentation.

Lees are the solid by-product of the production of Korean takju/ makgeolli rice wine. For our included study,[22] brown rice was soaked, steamed and cooled before being mixed with nuruk, a traditional fermentation starter. Nuruk is made by hanging a moistened wheat grain ‘cake’ to ferment for 2 to 4 weeks, attracting microorganisms such as *A. oryzae*, *R. oryzae,* LAB and yeasts.[23] Brown rice makgeolli was produced by fermenting and brewing this mixture. On completion, the separated lees underwent lyophilisation (removal of water and alcohol) to produce brown rice lees powder.[22] It is unclear which microorganisms remain in the brown rice lees.

Fermented papaya preparation is “made from the yeast fermentation of ripe pulp of Carica papaya using a specialized biofermentation technique”.[24]

Q-Can is a “patented food produced from organic soybeans which have been fermented with a guarded confidential mix of microorganisms and is available commercially as Q-Can Natural. The product is not a probiotic as it does not contain live cultures”.[25]

Micro-encapsulated fermented garlic extract is produced by the bacterial fermentation of decocted white garlic (*Allium sativum*). The cooled liquid garlic extract was fermented with a bacterial broth for 28 hours at 37°C, aged for one day at 80°C, filtered, heat concentrated and spray dried. For stability, carbohydrate-based food carriers were added. The resultant extract powder was then incorporated into dark chocolate.[26]
