## Supplemental material 4 for "The impact of botanical fermented foods on obesity, metabolic syndrome and type 2 diabetes: a systematic review of randomised controlled trials"

**Table: P-values of significant (p<0.05) between-group differences for each study**

| Parameter | Lipid | | | | | | | Glucose | | | | | | Anthropometric | | | | | | Blood pressure | | | Others | | | |
|---|---|---|---|---|---|---|---|---|---|---|---|---|---|---|---|---|---|---|---|---|---|---|---|---|---|---|
| **BFF (first author, year)** | TC | TG | LDL-C | HDL-C | ApoB | ApoA1 | hsCRP | FBG | HbA1c | HOMA-IR | FBI | 2hPPG | 2hPPI | BFP | BFM | BW | WC | WHR | VFA | DBP | SBP | n BP | CRP | ALT | AST | GM |
| *LACTOFERMENTED FOODS Lactobacillus spp.* | | | | | | | | | | | | | | | | | | | | | | | | | | |
| Fermented kimchi (Kim 2011) [1] | <0.05 | | | | | | | <0.05 | | | | | | <0.05 | | | | | | <0.05 | <0.05 | | | | | |
| Fermented kimchi (An 2012) [2] | No significant differences found between groups. | | | | | | | | | | | | | | | | | | | | | | | | | |
| Fermented kimchi (An 2013) [3] | No significant differences found between groups. | | | | | | | | | | | | | | | | | | | | | | | | | |
| Fermented kimchi (Lee 2013) [4] | | | | | | | | | | | | | | | | | | | | <0.05 | | | <0.05 | | | |
| Fermented red ginseng (Oh 2014) [5] | | | | | | | | | | | | 0.01 | 0.04 | | | | | | | | | | | | | |
| Fermented kimchi (Han 2015) [6] | No significant differences found between groups. | | | | | | | | | | | | | | | | | | | | | | | | | |
| Lactofermented Annurca apple (Tenore 2019) [7] | | | | <0.05 | | | | | | | | | | | | | | | | | | | | | | <0.05 |
| Ash kardeh (Salehi 2022) [8] | 0.03 | 0.00 | | 0.05 | | | | 0.00 | | | | | | | | | | | | | <0.00 | | | | | |
| *JANGS Bacillus & Aspergillus spp.* | | | | | | | | | | | | | | | | | | | | | | | | | | |
| Chungkookjang (Back 2011) [9] | 0.03 | | | | | | | | | | | | | | | | | | | | | | | | | |
| Red ginseng chungkookjang (Shin 2011) [10] | | | | | | | | <0.05 | | | | | | | | | | | | | | | | | | |
| Doenjang (Cha 2012) [11] | | | | | | | | | | | | | | 0.01 | <0.00 | <0.00 | | | 0.04 | | | | | | | |
| Kochujang (Cha 2013) [12] | | 0.1 | | | | | | | | | | | | | | | | | 0.04 | | | | | | | |
| Chungkookjang (Byun 2016) [13] | | | | | | | | | | | | | | 0.05 | | | 0 | 0.01 | | | | | | | | |
| Kochujang (Han 2022) [14] | No between-group analysis performed. | | | | | | | | | | | | | | | | | | | | | | | | | |
| *SHIOKOJI/ MISO/ KOCHUJANG/ AMAZAKE/ TOUCHI Aspergillus oryzae* | | | | | | | | | | | | | | | | | | | | | | | | | | |
| Touchi-extract houji tea (Fujita 2001) [15] | | | | | | | | <0.05 | <0.01 | | | | | | | | | | | | | | | | | |
| *A. oryzae* -fermented kochujang (Lim 2015) [16] | <0.05 | | | | | | | | | | | | | | | | | | | | | | | | | |
| Shiokoji (Nakamura 2020) [17] | No significant differences found between groups. | | | | | | | | | | | | | | | | | | | | | | | | | |
| Miso (Kondo 2019) [18] | | | | | | | | | | | | | | | | <0.05 | | | | | | <0.05 | | | | |
| Brown rice amazake (Akamine 2022) [19] | | | | | | | | | | | | | | | | | | | | | | | | | | <0.05 |
| *TEMPEH Rhizopus spp.* | | | | | | | | | | | | | | | | | | | | | | | | | | |
| Tempeh drink (Wirawanti 2017) [20] | | | 0.01 | | | | | | | | | | | | | | | | | | | | | | | |
| Tempeh gembus (Afifah 2020) [21] | | | 0.04 | | | | | | | | | | | | | | | | | | | | | | | |
| Tempeh (Nadia 2020) [22] | | 0 | | 0.00 | | | 0.03 | 0.00 | | | 0.00 | | | | | | | | | | | | | | | |
| *OTHERS* | | | | | | | | | | | | | | | | | | | | | | | | | | |
| Brown rice lees (Kim 2011) [23] | <0.05 | | <0.05 | | | | | | <0.05 | | | | | | | | <0.05 | | | | | | | <0.05 | <0.05 | |
| Fermented papaya preparation (Somanah 2012) [24] | | | | | | | | | | | | | | | | | | | | | | | 0.02 | | | |
| Fermented soy powder (Jung 2021) [25] | 0.00 | | 0.03 | 0.00 | | 0.04 | | | | | | | | | | | | | | | | | | | | |
| Fermented garlic chocolates (Mohammadi 2022) [26] | No significant differences found between groups. | | | | | | | | | | | | | | | | | | | | | | | | | |

TC: total cholesterol; TG: triglycerides; LDL-C: low density lipoprotein cholesterol; HDL-C: high density lipoprotein cholesterol; ApoB: apolipoprotein B; ApoA1: apolipoprotein A1; hsCRP: high sensitivity C-reactive protein; FBG: fasting blood glucose HbA1c: haemoglobin A1c; HOMA-IR: homeostatic model assessment of insulin resistance; FBI: fasting blood insulin; 2hPPG: 2-hour postprandial glucose; 2hPPI: 2-hour postprandial insulin; BFP: body fat percentage; BFM: body fat mass; BW: body weight; WC: waist circumference; WHR: waist-hip ratio; VFA: visceral fat area; DBP: diastolic blood pressure; SBP: systolic blood pressure; nBP: nighttime blood pressure; CRP: C-reactive protein; ALT: alanine transaminase; AST: aspartate aminotransferase; GGT: gamma-glutamyl transferase; GM: gut microbiota composition.

1 Kim EK, An SY, Lee MS, et al. Fermented kimchi reduces body weight and improves metabolic parameters in overweight and obese patients. Nutr Res 2011;31:436–43. doi:10.1016/j.nutres.2011.05.011

2 An SY, Lee MS, Choi YJ, et al. Favorable effects of kimchi on metabolic parameters and cytokine levels in patients with type 2 diabetes mellitus. Diabetes 2012;61:A190. doi:10.2337/db12-656-835

3 An SY, Lee MS, Jeon JY, et al. Beneficial effects of Kimchi on glucose metabolism related parameters in subjects with prediabetes. Diabetes 2013;62:A189. doi:10.2337/db13-680-858

4 Lee MS, An S.-Y., Jeon J.Y., et al. Beneficial effects of Kimchi on metabolic parameters in subjects with metabolic syndrome. Diabetes 2013;62:A191. doi:10.2337/db13-680-858

5 Oh M-R, Park S-H, Kim S-Y, et al. Postprandial glucose-lowering effects of fermented red ginseng in subjects with impaired fasting glucose or type 2 diabetes: a randomized, double-blind, placebo-controlled clinical trial. BMC Complement Altern Med 2014;14:237. doi:10.1186/1472-6882-14-237

6 Han K, Bose S, Wang J, et al. Contrasting effects of fresh and fermented kimchi consumption on gut microbiota composition and gene expression related to metabolic syndrome in obese Korean women. Mol Nutr Food Res 2015;59:1004–8. doi:10.1002/mnfr.201400780

7 Tenore GC, Caruso D, Buonomo G, et al. Lactofermented Annurca apple puree as a functional food indicated for the control of plasma lipid and oxidative amine levels: Results from a randomised clinical trial. Nutrients 2019;11:122. doi:10.3390/nu11010122

8 Salehi SO, Karimpour F, Imani H, et al. Effects of an Iranian traditional fermented food consumption on blood glucose, blood pressure, and lipid profile in type 2 diabetes: a randomized controlled clinical trial. Eur J Nutr 2022;61:3367–75. doi:10.1007/s00394-022-02867-2

9 Back H-I, Kim S-R, Yang J-A, et al. Effects of chungkookjang supplementation on obesity and atherosclerotic indices in overweight/obese subjects: A 12-week, randomized, double-blind, placebo-controlled clinical trial. J Med Food 2011;14:532–7. doi:10.1089/jmf.2010.1199

10 Shin S-K, Kwon J-H, Jeong Y-J, et al. Supplementation of cheonggukjang and red ginseng cheonggukjang can improve plasma lipid profile and fasting blood glucose concentration in subjects with impaired fasting glucose. J Med Food 2011;14:108–13. doi:10.1089/jmf.2009.1366

11 Cha Y-S, Yang J-A, Back H-I, et al. Visceral fat and body weight are reduced in overweight adults by the supplementation of Doenjang, a fermented soybean paste. Nutr Res Pract 2012;6:520–6. doi:10.4162/nrp.2012.6.6.520

12 Cha Y-S, Kim S-R, Yang J-A, et al. Kochujang, fermented soybean-based red pepper paste, decreases visceral fat and improves blood lipid profiles in overweight adults. Nutr Metab 2013;10:24. doi:10.1186/1743-7075-10-24

13 Byun M-S, Yu O-K, Cha Y-S, et al. Korean traditional Chungkookjang improves body composition, lipid profiles and atherogenic indices in overweight/obese subjects: a double-blind,

randomized, crossover, placebo-controlled clinical trial. Eur J Clin Nutr 2016;70:1116–22. doi:10.1038/ejcn.2016.77

14 Han AL, Jeong SJ, Ryu MS, et al. Anti-Obesity Effects of Traditional and Commercial Kochujang in Overweight and Obese Adults: a Randomized Controlled Trial. Nutrients 2022;14. doi:10.3390/nu14142783

15 Fujita H, Yamagami T, Ohshima K. Long-Term Ingestion of a Fermented Soybean-Derived Touchi-Extract with α-Glucosidase Inhibitory Activity Is Safe and Effective in Humans with Borderline and Mild Type-2 Diabetes. J Nutr 2001;131:2105–8. doi:10.1093/jn/131.8.2105

16 Lim J-H, Jung E-S, Choi E-K, et al. Supplementation with Aspergillus oryzae-fermented kochujang lowers serum cholesterol in subjects with hyperlipidemia. Clin Nutr 2015;34:383–7. doi:10.1016/j.clnu.2014.05.013

17 Nakamura A, Kitagawa M, Yamamoto T, et al. Fasting blood glucose-lowering effects of salted rice koji (Shiokoji) in mildly hyperglycemic adults - a randomized, double-blind, placebo-controlled parallel-group study. Jpn Pharmacol Ther 2020;48:215–24.

18 Kondo T, Kishi M, Fushimi T, et al. Vinegar Intake Reduces Body Weight, Body Fat Mass, and Serum Triglyceride Levels in Obese Japanese Subjects. Biosci Biotechnol Biochem 2009;73:1837–43. doi:10.1271/bbb.90231

19 Akamine Y, Millman JF, Uema T, et al. Fermented brown rice beverage distinctively modulates the gut microbiota in Okinawans with metabolic syndrome: A randomized controlled trial. Nutr Res 2022;103:68–81. doi:10.1016/j.nutres.2022.03.013

20 Wirawanti IW, Hardinsyah H, Briawan D, et al. Efek intervensi minuman tempe terhadap penurunan kadar low density lipoprotein. J Gizi Dan Pangan 2017;12:9–16. doi:10.25182/jgp.2017.12.1.9-16

21 Afifah DN, Nabilah N, Supraba GT, et al. The effects of tempeh gembus, an Indonesian fermented food, on lipid profiles in women with hyperlipidemia. Curr Nutr Food Sci 2020;16:56–64.

22 Nadia FS, Wati DA, Isnawati M, et al. The effect of processed tempeh gembus to triglycerides levels and insulin resistance status in women with obesity. Food Res 2020;4:1000-1010. doi:10.26656/fr.2017.4(4).415

23 Kim TH, Kim EK, Lee M-S, et al. Intake of brown rice lees reduces waist circumference and improves metabolic parameters in type 2 diabetes. Nutr Res 2011;31:131–8. doi:10.1016/j.nutres.2011.01.010

24 Somanah J, Bourdon E, Rondeau P, et al. Relationship between fermented papaya preparation supplementation, erythrocyte integrity and antioxidant status in pre-diabetics. Food Chem Toxicol 2014;65:12–7. doi:10.1016/j.fct.2013.11.050

25 Jung SM, Haddad EH, Kaur A, et al. A non-probiotic fermented soy product reduces total and LDL cholesterol: A randomized controlled crossover trial. Nutrients 2021;13:535. doi:10.3390/nu13020535

26 Mohammadi S, Mazloomi SM, Niakousari M, et al. Evaluating the effects of dark chocolate formulated with micro-encapsulated fermented garlic extract on cardio-metabolic indices in hypertensive patients: A crossover, triple-blind placebo-controlled randomized clinical trial. Phytother Res 2022;36:1785–96. doi:10.1002/ptr.7421
